# COVID-19 patients share common, corticosteroid-independent features of impaired host immunity to pathogenic molds

**DOI:** 10.1101/2022.04.21.22274082

**Authors:** Beeke Tappe, Chris D. Lauruschkat, Lea Strobel, Jezreel Pantaleón García, Oliver Kurzai, Silke Rebhan, Sabrina Kraus, Elena Pfeuffer-Jovic, Lydia Bussemer, Lotte Possler, Matthias Held, Kerstin Hünniger, Olaf Kniemeyer, Sascha Schäuble, Axel A. Brakhage, Gianni Panagiotou, P. Lewis White, Hermann Einsele, Jürgen Löffler, Sebastian Wurster

## Abstract

Patients suffering from coronavirus disease-2019 (COVID-19) are at high risk for deadly secondary fungal infections such as COVID-19-associated pulmonary aspergillosis (CAPA) and COVID-19-associated mucormycosis (CAM). Despite this clinical observation, direct experimental evidence for severe acute respiratory syndrome coronavirus type 2 (SARS-CoV-2)-driven alterations of antifungal immunity is scarce. Using an *ex-vivo* whole blood (WB) stimulation assay, we challenged blood from twelve COVID-19 patients with *Aspergillus fumigatus* and *Rhizopus arrhizus* antigens and studied the expression of activation, maturation, and exhaustion markers, as well as cytokine secretion. Compared to healthy controls, T-helper cells from COVID-19 patients displayed increased expression levels of the exhaustion marker PD-1 and weakened *A. fumigatus*- and *R. arrhizus*-induced activation. While baseline secretion of proinflammatory cytokines was massively elevated, WB from COVID-19 patients elicited diminished release of T-cellular (e.g., IFN-γ, IL-2) and innate immune cell-derived (e.g., CXCL9, CXCL10) cytokines in response to *A. fumigatus* and *R. arrhizus* antigens. Additionally, samples from COVID-19 patients showed deficient granulocyte activation by mold antigens and reduced fungal killing capacity of neutrophils. These features of weakened anti-mold immune responses were largely decoupled from COVID-19 severity, the time elapsed since diagnosis of COVID-19, and recent corticosteroid uptake, suggesting that impaired anti-mold defense is a common denominator of the underlying SARS-CoV-2 infection. Taken together, these results expand our understanding of the immune predisposition to post-viral mold infections and could inform future studies of immunotherapeutic strategies to prevent and treat fungal superinfections in COVID-19 patients.

## Introduction

Patients suffering from coronavirus disease 2019 (COVID-19) caused by severe acute respiratory syndrome coronavirus type 2 (SARS-CoV-2) are highly susceptible to fungal superinfections, especially COVID-19-associated pulmonary aspergillosis (CAPA) and COVID-19-associated mucormycosis (CAM) [1, 2]. CAPA has been encountered in approximately 15% of COVID-19 patients admitted to the intensive care unit (ICU), with regional high incidence rates of up to 39%, depending on classification criteria and diagnostic strategy [1, 3]. Similarly, devastating regional outbreaks of CAM considerably aggravated the death toll of COVID-19 and its burden on healthcare systems, especially during the spring 2021 wave of COVID-19 in India [2]. Deeper knowledge of the immunological factors driving the predisposition to CAPA and CAM will be essential to guide improved strategies for the prevention, targeted diagnosis, and treatment of these deadly coinfections.

Current hypotheses regarding the emergence of CAPA and CAM involve a combination of virus-induced damage of airway epithelia, severe immune dysregulation with impaired cellular immunity and exuberant hyperinflammatory cytokine release, and predisposing host factors such as structural lung diseases or poorly controlled diabetes mellitus [2-8]. These intrinsic risk factors are further compounded by iatrogenic immunosuppression (glucocorticosteroids [GCS] and/or monoclonal antibodies) and disruption of the microbiome by broad-spectrum antibiotics [2-8]. While the clinical risk factors predisposing COVID-19 patients to mold infections have been studied [2, 6], experimental evidence for impairment of anti-mold immunity resulting from the underlying viral infection is scarce. Instead, most of the presumed immunopathological mechanisms underlying the predisposition to CAPA and CAM were either extrapolated from other viral/fungal co-infections such as influenza-associated pulmonary aspergillosis (IAPA) or from global changes to the immune landscape in COVID-19 patients [9]. Furthermore, while the vastly different epidemiology of CAPA and CAM has been linked to underlying host factors (e.g., the high number of patients with unknown or uncontrolled diabetes mellitus in India) [6, 10, 11], and, possibly, environmental exposures [10], little is known about similarities or differences in SARS-CoV-2-driven impairment of immune responses to *Aspergillus* and Mucorales.

Therefore, we herein studied mold antigen-induced immune responses in a cohort of COVID-19 patients without classical host factors for mold infections (e.g., active hematological malignancies and prolonged neutropenia). We found several conserved features of diminished antifungal host defense in patients with COVID-19, including signs of increased T-helper (Th) cell exhaustion, impairment of mold-induced cytokine release, and neutrophil dysfunction with diminished fungicidal activity. These alterations were largely unaffected by the causative SARS-CoV-2 variant, GCS treatment, infection severity, and the time elapsed since diagnosis of COVID-19, suggesting a generic impairment of anti-mold defense in patients with SARS-CoV-2 infections.

## Materials and Methods

### Ethics statement

This study was approved by the Ethics Committee of the University of Würzburg (protocol number 152/20). Informed written consent was obtained from all participants.

### Subjects

The patient cohort consisted of adult patients (≥ 18 years of age) treated for COVID-19 at University Hospital of Würzburg, Missionsärztliche Klinik Würzburg, or Main-Klinik Ochsenfurt (all in Bavaria, Germany) between March and December 2021. The diagnosis of COVID-19, defined as the time of the first positive SARS-CoV-2 polymerase chain reaction (PCR) on a nasopharyngeal or oropharyngeal sample, had to be within 4 weeks prior to blood collection. All patients had to have moderate disease severity at the time of blood collection, i.e., a WHO progression scale (WHO score) of 4 or 5 [12]. Exclusion criteria were pregnancy, known coinfections, vaccinations within the past 4 weeks or any prior SARS-CoV-2 vaccination, chronic infectious diseases, active cancer, and any antifungal or immunomodulatory therapy within the past 12 weeks, except for GCS initiated for the treatment of COVID-19. The control cohort consisted of healthy adults without prior SARS-CoV-2 infection or vaccination, with the same exclusion criteria as for the patient cohort.

### Clinical chart review

The following clinical parameters were recorded: Age, gender, time elapsed since the first positive SARS-CoV-2 PCR, respiratory support, highest WHO score until blood collection, GCS therapy (agent, dose, route of administration), as well as preexisting cardiovascular, metabolic, and pulmonary diseases. In addition, blood count and clinical chemistry at the time of immune cell sampling were reviewed.

### Blood collection

Venous blood was collected using the Monovette^®^ blood collection system (Sarstedt, Nürnbrecht, Germany). Lithium-heparin- and hirudin-anticoagulated blood was used for whole blood (WB)-based immunoassays. Polymorphonuclear cells (PMNs) were isolated from EDTA-anticoagulated blood. All blood samples were processed within 2 h of collection.

### Preparation of fungal lysates

A protein extract from an *Aspergillus fumigatus* mycelial lysate (AfuLy) was obtained as described previously [13]. A *Rhizopus arrhizus* lysate (RarLy) was obtained from an in-house clinical isolate (Rar-2021-10, Institute of Hygiene and Microbiology, Würzburg). Spores were incubated in Roswell Park Memorial Institute medium (RPMI, Gibco, Thermo Fisher Scientific, Waltham, MA, USA; 1×10^6^ spores/mL) at 25 °C in a shaking incubator (200 rpm) until mycelial clusters were visible. Hyphal suspensions were centrifuged at 5000×*g* for 10 min and supernatants were discarded. Mycelium was washed with Hank’s Balanced Salt Solution (HBSS, Sigma-Aldrich, St. Louis, MO, USA) and homogenized in 1 mL HBSS using a NucleoSpin Bead Tube Type A (Macherey-Nagel, Düren, Germany) and a Vortex-Genie 2 vortex mixer (Scientific Industries, Bohemia, NY, USA). Lysates were filter-sterilized using a 0.2-µm filter (Miltenyi Biotec, Bergisch Gladbach, Germany). Protein concentrations were determined with the DC Protein Assay Kit (Bio-Rad, Hercules, CA, USA) following the manufacturer’s instructions. Absorption was read at 750 nm in a microplate reader (Tecan, Maennerdorf, Switzerland). Endotoxin levels of the lysate were <0.3 endotoxin units per mg protein, as determined with the Pierce™ LAL Chromogenic Endotoxin Quantitation Kit (Thermo Fisher Scientific, Waltham, MA, USA) according to the manufacturer’s instructions.

### Preparation of germlings for stimulation of innate immune cells

*A. fumigatus* strain American Type Culture Collection (ATCC) 46645 was plated on beer wort agar (Oxoid, Wesel, Germany) and incubated at 30 °C for 4 days. The plate was rinsed with sterile distilled water to harvest the conidia and the conidial suspension was passed through a 20-µm cell strainer (Miltenyi Biotec, Bergisch Gladbach, Germany). To generate *A. fumigatus* germlings (AfuG), RPMI medium (20 mL in 50-mL tubes) was inoculated with 2×10^7^ conidia. Tubes were incubated at 25 °C and 200 rpm until germlings reached a length of 10-30 µm. After centrifugation at 5000×*g* for 10 min, germlings were resuspended in RPMI medium at 1×10^6^ AfuG/mL.

*R. arrhizus* (Rar-2021-10) was plated on beer wort agar and incubated at 30 °C for 6 days. Spores were harvested as described above. *R. arrhizus* germlings (RarG, length 20-40 µm) were generated at 37 °C in RPMI medium without shaking.

### *Ex-vivo* WB stimulation for immunoassays

To analyze adaptive immunity and cytokine release, *ex-vivo* WB stimulation was performed in 2.7-mL Monovette^®^ tubes, as previously described [13]. Briefly, tubes without anticoagulants were prepared with antigenic stimuli (AfuLy, RarLy, and/or SARS-CoV-2 Protein S [PrS, Miltenyi Biotec, Bergisch Gladbach, Germany]), co-stimulatory antibodies (α-CD28 and α-CD49d, Miltenyi Biotec, Bergisch Gladbach, Germany), and RPMI medium, as summarized in **Table S1**. Phytohemagglutinin (Roche, Basel, Switzerland) without co-stimulatory antibodies was used as a positive control. Tubes were cryopreserved at -20 °C for up to 4 weeks. Tubes were thawed, kept at 37 °C for at least 30 min prior to blood injection, and thoroughly disinfected with ethanol. Five-hundred microliters of heparinized WB was injected with an insulin syringe. Tubes were then incubated for 24 h at 37 °C. For flow cytometric analyses, brefeldin A (10 µg/mL, Sigma-Aldrich St.Louis, MO, USA) was added after 4 h of incubation.

For innate immune cell stimulation, a modified WB stimulation approach was used [14]. Tubes were prepared as described above and as summarized in **Table S1**. Differences compared to WB stimulation for adaptive immunoassays were i) omission of co-stimulatory antibodies, ii) the use of germlings (AfuG, RarG) instead of lysates, iii) the use of hirudin-anticoagulated WB instead of heparinized WB, and iv) a shorter incubation period of 4 h at 37 °C.

### Multiplex cytokine assays

WB was stimulated as described above. Tubes were centrifuged at 2000×*g* for 20 min. Plasma was collected, aliquoted, and cryopreserved at -80 °C until further analysis. Cytokine and chemokine concentrations were measured using the Cytokine Human Magnetic 35-Plex Panel (Thermo Fisher Scientific, Waltham, MA, USA) according to the manufacturer’s instructions. Acquisition was performed using a Luminex detection system (Bio-Plex 200 system) and Bio-Plex Manager(tm) Software 6.2 (Bio-Rad, Hercules, CA, USA).

### Flow cytometry

For the analysis of innate immunity and baseline characterization of leukocyte distributions, WB samples were centrifuged for 5 min at 2000×*g* at room temperature (RT). Cells were resuspended in 500 µL HBSS and aliquoted for different antibody panels. Antibodies for extracellular staining were added as detailed in **Table S2**, along with the fixable Viobility™ Live/Dead Dye (Miltenyi Biotec, Bergisch Gladbach, Germany) and dihydrorhodamine 123 (Sigma-Aldrich, St. Louis, MO, USA) for visualization of reactive oxygen species (ROS). After incubation in the dark for 20 min at RT, erythrocyte lysis buffer (EL buffer, Qiagen Inc., Venlo, Netherlands) was added. Tubes were incubated for 2 min (with repeated inversion) and centrifuged at 5000×*g* for 5 min. Supernatants were discarded and the erythrocyte lysis step was repeated. Cells were washed with 1 mL HBSS, resuspended in 200 µL of 4% paraformaldehyde (Sigma-Aldrich, St. Louis, MO, USA), incubated for 30 min at RT, and acquired on a MACS Quant 10 flow cytometer (Miltenyi Biotec, Bergisch Gladbach, Deutschland).

For flow cytometric assessment of T cells, WB was processed and stained as described before [13]. Antibody panels are summarized in **Table S2**. Samples were acquired on a Cytoflex AS34240 flow cytometer (Beckman Coulter, Brea, CA, USA).

Downstream data analysis was performed with Kaluza v.2.1 (Beckman Coulter, Brea, CA, USA). Gating strategies and representative raw data are shown in **Figure S1** and **S2** (adaptive immunity panels) and **Figure S3** (innate immunity panels).

### Fungicidal activity of polymorphonuclear cells

*A. fumigatus* and *R. arrhizus* conidia (2.5×10^5^ conidia/spores per well, same strains as used for the lysates) were seeded in 48-well plates in 250 µL of colorless RPMI medium supplemented with 10% fetal calf serum (FCS, Sigma-Aldrich, St. Louis, MO, USA) and incubated until germination.

To isolate PMNs, EDTA-anticoagulated blood was layered on a polysaccharide gradient (Polymorphprep, ProteoGenix, Schiltigheim, France) and centrifuged at 590×*g* for 30 min. The PMN interphase was harvested, pelleted for 5 min at 300×*g*, and resuspended in EL buffer to lyse the remaining erythrocytes. PMNs were washed with 50 mL of HBSS, centrifuged, counted with a hemocytometer, and resuspended in RPMI + 10% FCS. Aliquots of 5×10^4^, 1.25×10^5^, and 2.5×10^5^ PMNs in 250 µL RPMI + 10% FCS were added to the germlings to obtain effector/target (E:T) ratios of 0.2, 0.5, and 1. Blank wells (medium only) and wells containing either only PMNs or only germ tubes were used as controls. *A. fumigatus*/PMN cocultures and *R. arrhizus*/PMN cocultures were incubated for 2 h and 4 h, respectively, at 37 °C with 5% CO_2_. Optimal coculture periods and E:T ratios were determined in preceding experiments.

After incubation, wells were washed twice with 1 mL of ice-cold water for hypotonic lysis of PMNs (5 min incubation period per wash step). Hyphae were resuspended in 100 µL prewarmed RPMI + 10% FCS. Two-hundred microliters of prewarmed HBSS supplemented with 400 µg/mL of 2,3-bis-(2-methoxy-4-nitro-5-sulphenyl)-(2H)-tetrazolium-5-carboxanilide (XTT, Alfa Aesar, Thermo Fisher Scientific, Waltham, MA, USA) and 50 µg/mL of 2,3-dimethoxy-5-methyl-p-benzoquinone-coenzyme (Sigma-Aldrich, St. Louis, MO, USA) were added. After 45 min of incubation at 37 °C, triplicates of 100 µL supernatant were transferred to a 96-well plate and OD_450_ was measured in a microplate reader (Tecan, Maennerdorf, Switzerland). Fungal XTT metabolism was calculated according to the following formula: 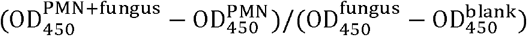.

### Statistics

Data compilation, statistical analyses, and visualization were performed using Microsoft Excel and GraphPad Prism v.9 (GraphPad Software, San Diego, CA, USA). Median-to-median ratio (MMR) was used as a descriptive surrogate of effect sizes between patients and controls. The 2-tailed Mann-Whitney U test (patients versus controls) and 2-tailed paired Wilcoxon test (paired samples from healthy controls) were used for significance testing, with subsequent Benjamini-Hochberg test for a false-positive discovery rate (FDR) of < 0.2. Spearman’s rank correlation coefficients (two continuous variables) and rank-biserial correlation coefficients (continuous variable versus dichotomous variable) were used for correlation analyses. Principal component analysis (PCA) of cytokine profiles was performed with ClustVis (https://biit.cs.ut.ee/clustvis/) [15] based on ln(x)+1-transformed data.

### Enrichment analysis

Signatures of baseline and antigen-induced expression levels of activation/maturation/exhaustion markers and logarithmically-transformed cytokine concentrations were analyzed using QIAGEN IPA core analysis (digitalinsights.quiagen.com, Qiagen Inc., Venlo, Netherlands) [16] to determine canonical pathway enrichment. Mean-based expression ratios between COVID-19 patients and controls were compared against the gene and chemical Ingenuity Knowledge Base modules, considering all confidence levels to identify direct and indirect relationships. Enrichments of canonical pathways were considered significant at an absolute z-score value ≥ 1 and a Benjamini-Hochberg adjusted p-value < 0.05.

## Results

Clinical characteristics of COVID-19 patients and baseline comparison of global immune features between patients and controls

Twelve COVID-19 patients were enrolled, four during the Alpha (B.1.1.7) wave and eight during the Delta (B.1.617.2) wave. Demographic and clinical characteristics are summarized in **Figure 1A**. Age distributions of patients (range 44-79, median 60.5) and healthy controls (n = 9, range 33-70, median 62) were comparable (p = 0.767). All patients required oxygen support at some point during their COVID-19 treatment, with 2 out of the 12 patients requiring invasive mechanical ventilation. Maximum WHO scores ranged from 5 to 9 (median 5). At the time of immune cell sampling (1-27 days after first positive SARS-CoV-2 PCR, median 13 days), all patients were hospitalized on regular wards and either required no oxygen support (WHO score 4) or received oxygen via nasal cannula (WHO score 5). Five patients had underlying pulmonary diseases (3 COPD, 1 asthma, 1 allergic bronchitis). Nine out of the 12 patients received GCS (all dexamethasone) as treatment for COVID-19 within the past 7 days, totaling to 0-280 mg prednisolone equivalent (median 120 mg).

**Figure 1.**
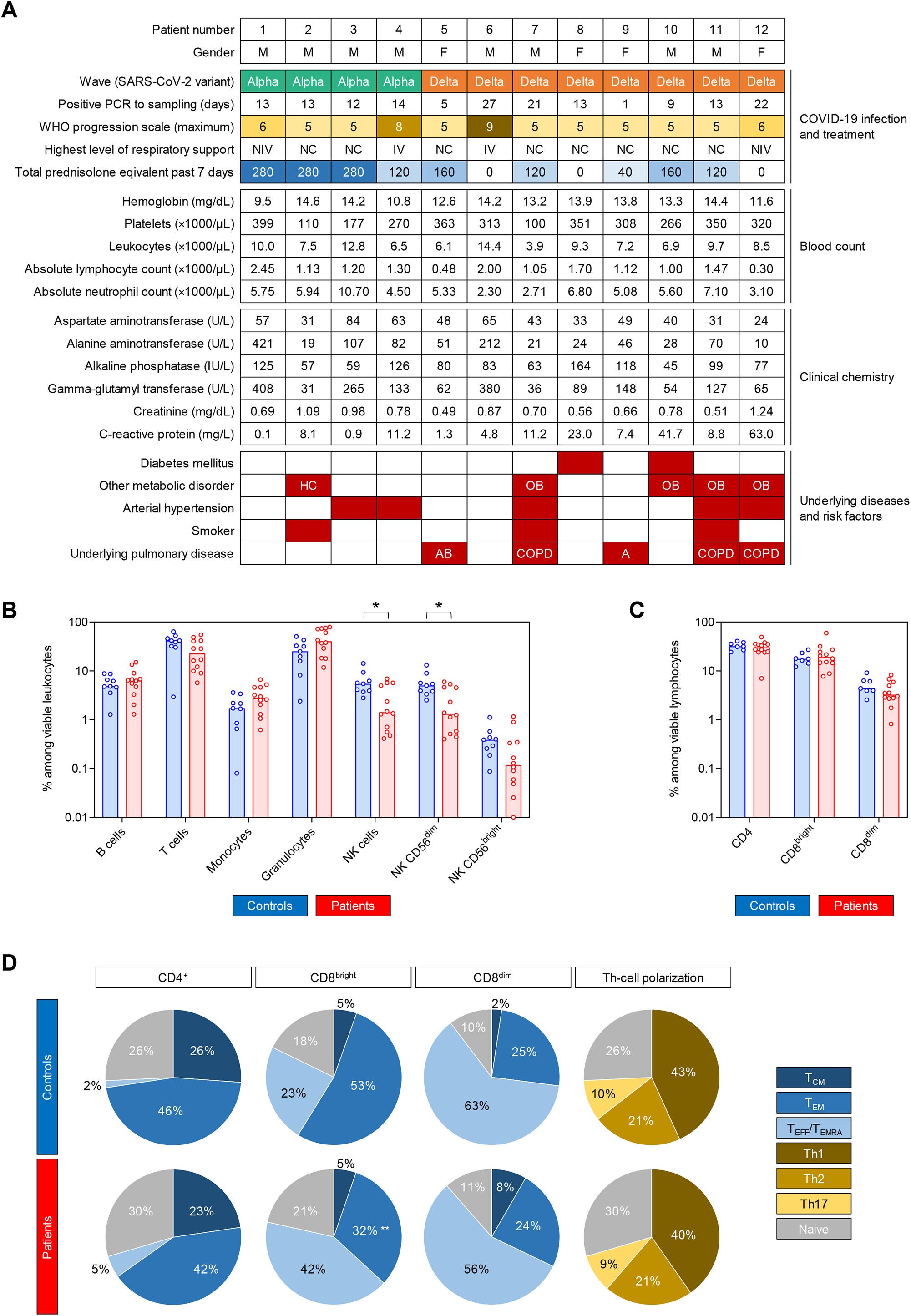
COVID-19 patients and control subjects have comparable global immune features and T-cell phenotypes. (**A**) Summary of clinical features and laboratory parameters of the patient cohort. (**B**) Percent distributions of cellular subsets among viable leukocytes in COVID-19 patients and controls. (**C**) Distributions of T-cell subsets among viable lymphocytes. (**B-C**) Individual values (dots) and medians (columns) are shown. (**D**) Mean distributions of memory/effector phenotypes among CD4^+^ T-helper (Th) cells, CD8^bright^ cytotoxic T lymphocytes (CTLs), and CD8^dim^ CTLs, as well as polarization of Th cells. (**B-D**) Mann-Whitney U test (patients versus controls) and Benjamini-Hochberg procedure to test for a false-positive discovery rate (FDR) of < 0.2. * p < 0.05, ** p < 0.01. <u>Abbreviations:</u> M = male, F = female, PCR = polymerase chain reaction, NC = nasal cannula, NIV = non-invasive ventilation, IV = invasive ventilation, (I)U = (international) unit, HC = hypercholesterinemia, OB = obesity, AB = allergic bronchitis, COPD = chronic obstructive pulmonary disease, A = asthma, T_CM_ = central memory T cells, T_EM_ = effector memory T cells, T_EFF_/T_EMRA_ = effector T cells/terminally differentiated effector memory T cells re-expressing CD45RA.

None of the patients was neutropenic at the time of immune cell sampling and only 2 out of the 12 patients had lymphocytopenia (defined as an absolute lymphocyte count < 1000/μL). Medians for leukocyte, neutrophil, and lymphocyte counts were 8000/μL, 5470/μL, and 1170/μL, respectively. C-reactive protein (CRP) values of 8 patients exceeded the normal range (0-5 mg/L) and five patients showed strongly elevated CRP (>10 mg/L). Median CRP was 8.46 mg/L.

The patient cohort had a more granulocyte-prone leukocyte distribution than control subjects (median 40.3% vs. 25.2%), with a trend toward lower relative T-cell portions among viable leukocytes (median 22.7% vs. 41.7%) and significantly lower natural killer (NK)-cell frequencies (median 1.4% vs. 5.5%, p = 0.019, **Figure 1B**). However, the median absolute NK-cell count (147/μL) in COVID-19 patients was well within the normal range and only two patients had NK-cell counts below 50/μL.

Distributions of T-helper (Th) cells and cytotoxic T-lymphocyte (CTL) populations among viable lymphocytes were normal and comparable between patients and controls (**Figure 1C**). Effector/memory phenotypes of Th cells and CTLs were similar in patients and controls, except for a shift from effector memory cells toward an effector/T_EMRA_ phenotype among CD8^bright^ CTLs (p = 0.005, **Figure 1D**) in COVID-19 patients, which is expectable during or after an acute viral infection [17, 18]. Th polarization was comparable in both cohorts, with type-1 Th cells (Th1) being the predominant phenotype (43% in controls, 40% in patients, **Figure 1D**). Although most patients had signs of acute inflammation (CRP, leukocytosis), these data suggest that COVID-19 patients and controls enrolled in this study had largely comparable baseline features of T-cell immunity.

### *A. fumigatus* antigen-reactive T-cell repertoire

In order to evaluate cellular activation and cytokine release in response to *A. fumigatus* antigens, we used a previously established WB-based stimulation system (**Figure 2A**) [13, 19]. Stimulation conditions were pre-optimized for each cell type (**Figure 2B**). Background-adjusted antigen-induced responses were defined as the difference between antigen-reactive response and unstimulated background (**Figure 2A**). For flow cytometric markers with high baseline expression (>25% of the cell population of interest), we additionally compared fold changes of mean fluorescence intensity (MFI).

**Figure 2.**
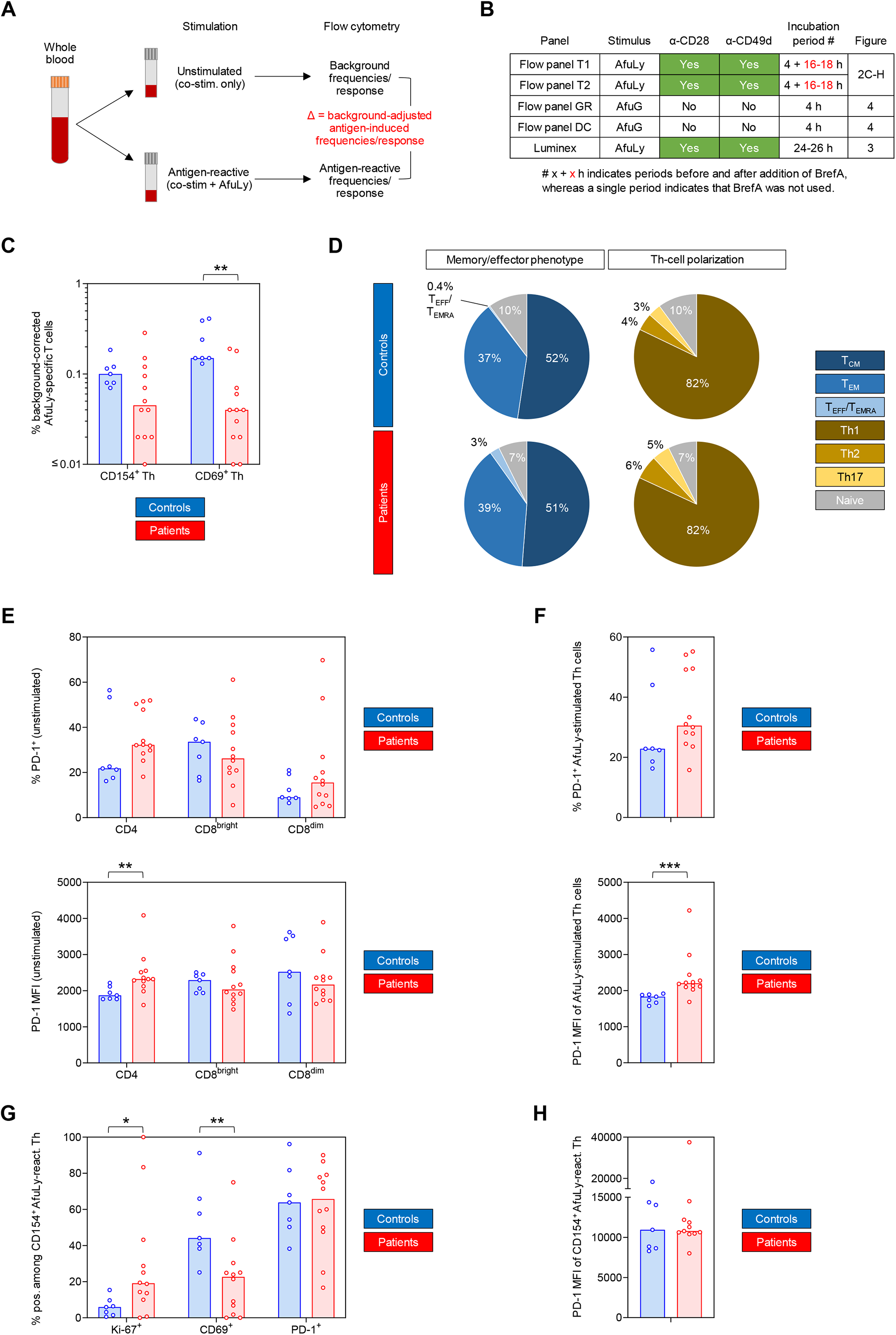
Whole blood-based T-cell phenotyping reveals impaired *Aspergillus* antigen-induced T-helper (Th) cell activation and increased Th exhaustion in samples from COVID-19 patients. (**A**) Assay principle and definitions of antigen-reactive and background-adjusted antigen-induced responses. (**B**) Summary of stimulation protocols. (**C**) Background-corrected frequencies of AfuLy-specific cells among CD4^+^ Th cells detectable by CD154 or CD69 upregulation. (**D**) Mean distributions of memory/effector phenotypes and subsets of AfuLy-reactive Th cells. (**E**) Frequencies of PD-1^+^ cells and PD-1 MFI among global T-cell subsets in unstimulated samples. (**F**) Frequencies of PD-1^+^ cells and PD-1 MFI among Th cells (all CD4^+^ cells) in AfuLy-stimulated samples. (**G**) Frequencies of CD69^+^, Ki-67^+^, and PD-1^+^ cells among CD4^+^ CD154^+^ AfuLy-reactive Th cells. (**H**) Mean fluorescence intensity (MFI) of PD-1 in CD4^+^ CD154^+^ AfuLy-reactive Th cells. (**C, E-H**) Columns represent medians. (**C-H**) Mann-Whitney U test (patients versus controls) and Benjamini-Hochberg procedure to test for a false-positive discovery rate (FDR) of < 0.2. * p < 0.05, ** p < 0.01, *** p < 0.001. <u>Abbreviations:</u> AfuLy = *Aspergillus fumigatus* lysate, AfuG = *Aspergillus fumigatus* germlings, co-stim. = co-stimulatory antibodies, BrefA = brefeldin A, PD-1 = programmed cell death protein 1, MFI = mean fluorescence intensity.

With regard to CD154 expression, the best-characterized flow cytometric marker for comprehensive mold-reactive Th-cell quantification [20, 21] and surrogate of recent and long-term mold exposure [22, 23], the patient cohort had lower background-corrected AfuLy-induced T-cell frequencies (median 0.05%) than controls (median 0.10%, **Figure 2C**). However, this trend was non-significant (p = 0.086) and both cohorts had generally rather low frequencies of CD154-responsive Th cells, suggesting that the results of the subsequent assays were not confounded by significant differences in prior mold encounters (e.g., through occupational exposures). In contrast, patients had significantly lower CD69^+^ background-corrected AfuLy-responsive T-cell frequencies than controls (median 0.04% vs. 0.15%, p = 0.002, **Figure 2C**), suggesting a potential deficit in Th activation. Memory/effector phenotypes and polarization of CD154^+^ AfuLy-reactive cells were comparable among patients and controls, with a predominance of central memory cells and Th1 cells (**Figure 2D**).

COVID-19 patients had higher PD-1 positive Th frequencies and higher PD-1 MFIs in both unstimulated (**Figure 2E**, MMR 1.24 for MFI fold changes, p = 0.005) and AfuLy-stimulated samples (**Figure 2F**, MMR 1.20 for MFI fold changes, p < 0.001). Cells responding to the lysate by CD154 upregulation showed strong PD-1 expression in both cohorts (**Figure 2G-H**), aligning with the known potential of *Aspergillus* antigens as drivers of T-cell exhaustion [24]. Interestingly, CD154^+^ AfuLy-reactive cells of COVID-19 patients showed stronger Ki-67 expression than controls (MMR 3.24, p = 0.028, **Figure 2G**), whereas baseline Ki-67 expression among all Th cells was comparable (p = 0.547, data not shown). This observation suggests an enrichment of highly proliferative AfuLy-reactive cells in the patient cohort, whereas less proliferative (and potentially exhausted) Th cells might have failed to sufficiently upregulate activation markers beyond the detection threshold. This hypothesis is supported by the CD69 expression deficit in Th cells from COVID-19 patients that persisted when restricting the analysis to CD154^+^ AfuLy-reactive cells (**Figure 2G**). In summary, these data suggest a modest increase in Th-cell exhaustion and a trend toward weakened Th activation in COVID-19 patients versus controls.

### Baseline and *A. fumigatus* antigen-induced cytokine responses

Next, we quantified a set of 35 cytokines and chemokines to analyze the AfuLy-induced cytokine release in WB samples from COVID-19 patients and controls. PCA plots suggested increasing segregation of the two cohorts from unstimulated background to AfuLy-reactive responses to background-corrected AfuLy-induced responses (**Figure 3A**). The latter comparison showed almost complete separation of 95% confidence areas between patients and controls, with one notable outlier in the patient cohort (highlighted with a # in **Figures 3A** and **B**).

**Figure 3.**
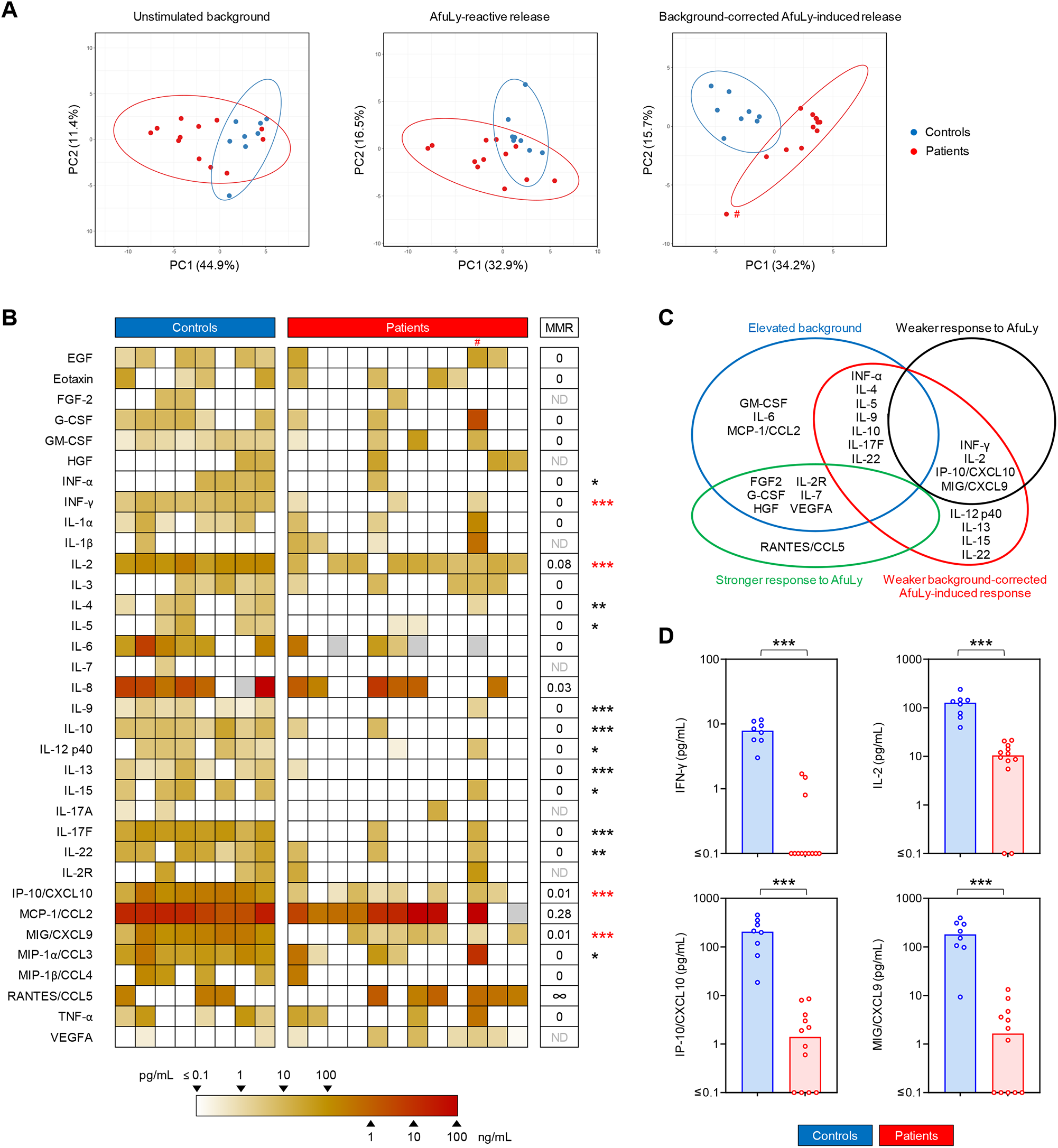
Whole blood from COVID-19 patients elicits weakened *Aspergillus fumigatus*-induced cytokine responses. (**A**) Principal component analysis (PCA) comparing unstimulated background secretion, *Aspergillus* lysate (AfuLy)-reactive, and background-adjusted AfuLy-induced cytokine release. Ellipses represent 95% confidence ranges. (**B**) Heat map representing individual background-adjusted AfuLy-induced cytokine responses. IL-1RA is not shown (0 pg/mL in all subjects). Grey boxes indicate non-determinable values (i.e., measurements with unstimulated background exceeding the detectable range). # denotes the outlier in the PCA. MMR = median-to-median ratio (patients/controls). ∞ = infinite MMR (median 0 pg/mL in the control cohort). ND = MMR not defined (median 0 pg/mL in both cohorts). Mann-Whitney U test (patients versus controls) and Benjamini-Hochberg procedure to test for a false-positive discovery rate (FDR) of < 0.2. * p < 0.05, ** p < 0.01, *** p < 0.001. (**C**) Venn diagram summarizing statistically significant alterations in unstimulated background, AfuLy-reactive, and background-adjusted AfuLy-induced cytokine responses in COVID-19 patients versus controls. Significance was defined as p < 0.05 and FDR < 0.2. (**D**) Individual and median (columns) background-adjusted AfuLy-induced concentrations of cytokines that showed both, significantly weaker AfuLy-reactive and background-adjusted AfuLy-induced release in COVID-19 patients, indicated by red asterisks in panel (**B**). *** p < 0.001.

Fifteen cytokines displayed significantly lower background-corrected AfuLy-induced concentrations in patients compared to controls (**Figure 3B**). Twelve out of these 15 cytokines had no detectable background-corrected AfuLy-induced release in the majority of patients (median 0 pg/mL), and the 3 remaining cytokines (IL-2, IP-10/CXCL10, MIG/CXCL9) had MMRs of <0.1 in patients versus controls (**Figure 3B**). T-cellular cytokines, including surrogates of Th1 (e.g., IFN-γ, IL-2) and Th2 (IL-4, IL-5, IL-13) activation, accounted for most of the significantly disparate responses. Considering the etiology of differential background-corrected AfuLy-induced release, that is, elevated unstimulated background (**Figure S4**) versus genuinely impaired AfuLy-reactive responses, a subset of four cytokines (IFN-γ, IL-2, IP-10/CXCL10, MIG/CXCL9) showed both significantly reduced AfuLy-reactive and background-corrected AfuLy-induced responses (**Figure 3C**, red asterisks in **Figure 3B**, individual concentrations shown in **Figure 3D**).

Based on prior experience with mutual impairment of immune responses during dual viral and *A. fumigatus* antigen challenge [25], we sought to test whether WB from healthy donors co-challenged with AfuLy and PrS can recapitulate the deficit in AfuLy-induced cytokine responses seen in COVID-19 patients. However, none of the 35 tested cytokines displayed significantly altered release upon dual antigen exposure compared to AfuLy alone (**Figure S5**), suggesting that impaired T-cellular cytokine release in samples from COVID-19 patients is predominantly driven by preexisting *in-vivo* alterations such as Th-cell exhaustion.

### *A. fumigatus*-induced activation and fungicidal activity of innate immune cells

Further, we utilized our *ex-vivo* WB stimulation assay to test the reactivity of innate immune cell subsets to *A. fumigatus*. Granulocytes from control subjects showed strong induction of ROS production (median MFI fold change 18.9), upregulation of CD62L (median MFI fold change 8.8), and modest induction of CD253 (median MFI fold change, 1.3) upon challenge with AfuG. All 3 responses were strongly and significantly impaired in granulocytes from COVID-19 patients, with median MFI fold changes of 2.8 for ROS (MMR vs. controls 0.15, p < 0.001), 2.7 for CD62L (MMR 0.31, p = 0.006), and 1.0 for CD253 (MMR 0.72, p = 0.010, **Figure 4A-B**). Significant differences in AfuG-induced granulocyte activation between patients and controls were confirmed when comparing activation marker-positive granulocyte frequencies instead of MFI (**Figure 4B**). Additionally, granulocytes from COVID-19 patients showed a trend toward lower baseline expression of the maturation marker CD16 in unstimulated samples (MMR 0.71, **Figure S6A**).

**Figure 4.**
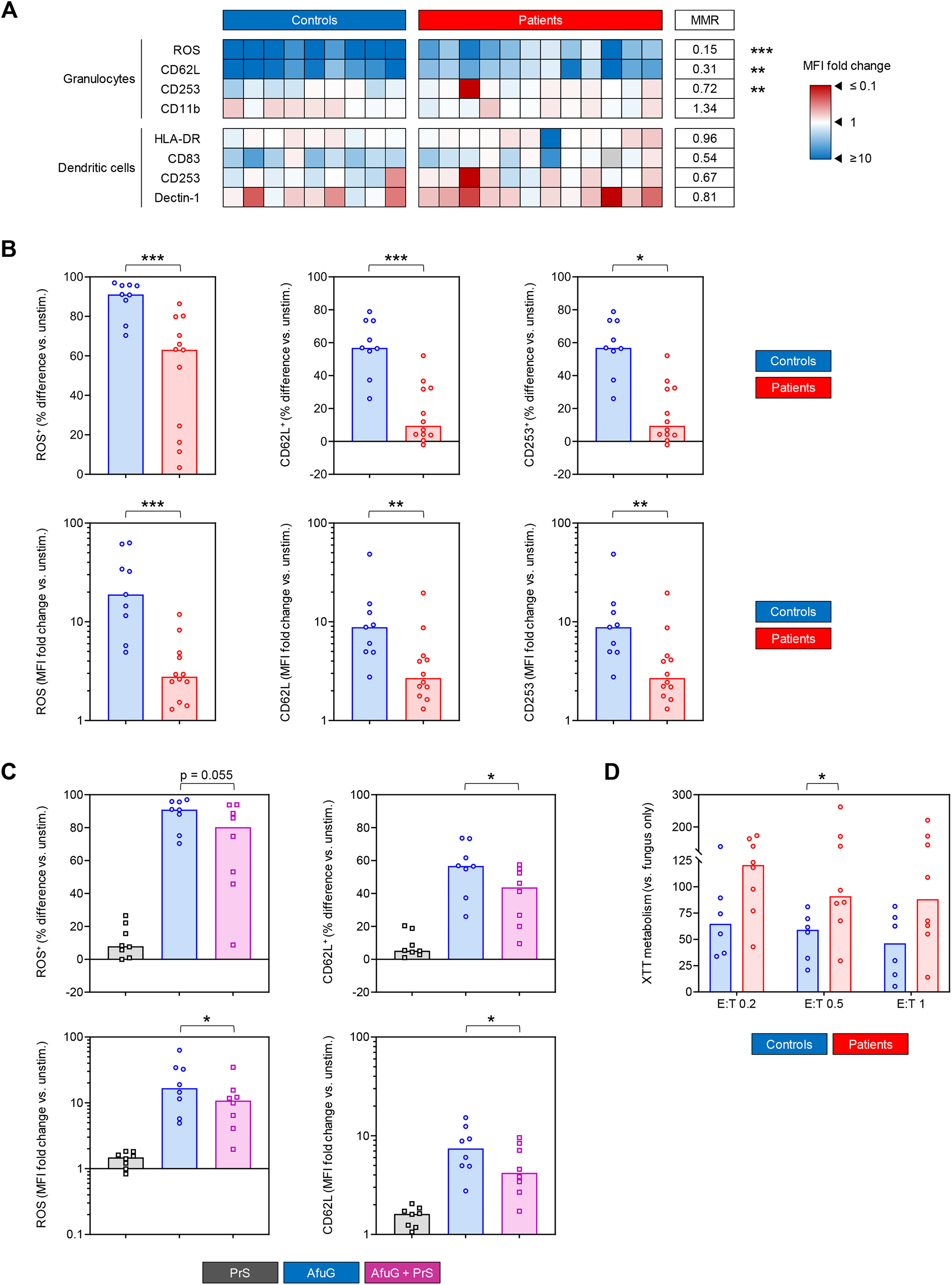
Granulocytes from COVID-19 patients show diminished *Aspergillus fumigatus*-induced activation and fungal killing potential. (**A**) Heat map summarizing induction of activation markers in/on granulocytes and dendritic cells (DCs) upon stimulation with *Aspergillus fumigatus* germlings (AfuG). Fold changes of mean fluorescence intensity (MFI) in stimulated versus unstimulated samples are represented by color scale. MMR = median-to-median ratio (patients/controls). Grey boxes indicate non-determinable ratios (i.e., measurements with a baseline MFI ≤ 0). (**B**) Individual and median (columns) differential frequencies and MFI fold changes of key activation markers in/on granulocytes in AfuG-stimulated whole blood (WB) versus unstimulated WB. (**C**) Individual and median (columns) frequencies and MFI fold changes of key activation markers in/on granulocytes in WB stimulated with AfuG, SARS-CoV2 Protein S (PrS), and a combination of AfuG + PrS. All responses are normalized to unstimulated control samples. (**D**) Relative hyphal proliferation in the presence of neutrophils at different effector/target (E:T) ratios, normalized to a “fungus only” control without neutrophils. Lower values indicate stronger killing potential of neutrophils. (**A-D**) Mann-Whitney U test (patients versus controls, **A, B, D**) or paired Wilcoxon test (paired samples from healthy controls, **C**) and Benjamini-Hochberg procedure to test for a false-positive discovery rate (FDR) of < 0.2. * p < 0.05, ** p < 0.01, *** p < 0.001.

Similarly, dendritic cells (DCs) from COVID-19 patients had weaker baseline expression of maturation markers CD83 (MMR 0.75) and HLA-DR (MMR 0.59, p = 0.018), along with a trend toward reduced expression of pattern recognition receptors (PRR) CD284/TLR4 (MMR 0.29) and Dectin-1 (MMR 0.62, **Figure S6B**). In contrast, we found no differences in AfuG-induced DC activation between patients and controls, except for a non-significant trend toward weaker upregulation of the maturation marker CD83 in patient samples (MMR 0.54, **Figure 4A**).

Unlike the deficient T-cellular cytokine responses, impaired granulocyte responses were partially reproducible when WB from healthy donors underwent concomitant stimulation with AfuG and PrS. While PrS induced slight activation of granulocytes from most healthy donors, dual challenge with AfuG + PrS weakened the fungus-induced response compared to AfuG alone (e.g., median ratio in fold changes of ROS MFI 0.49, p = 0.039, **Figure 4C**). However, changes in marker expression after dual antigen challenge versus AfuG alone were considerably weaker compared to differences between COVID-19 patients and controls. This observation suggests that both underlying *in-vivo* alterations (e.g., exhaustion) and immune attenuation during dual antigen exposure contribute to the observed post-COVID-19 granulocyte dysfunction.

Given the impaired granulocyte activation and ROS response, we hypothesized that neutrophils from COVID-19 patients might elicit less potent fungicidal activity. Testing this hypothesis in a metabolic fungal killing assay, we found stronger XTT metabolism by *A. fumigatus* germlings after coculture with PMNs from patients versus controls at all 3 E:T ratios tested (MMR 1.59-1.91, p = 0.029 at E:T 0.5, **Figure 4D**). Taken together, these findings indicate that granulocytes from COVID-19 patients have weakened reactivity to *A. fumigatus* antigens and impaired fungicidal activity.

### Impact of causative SARS-CoV-2 variant and clinical characteristics on surrogates of anti-*Aspergillus* immunity

In view of the known differences in host-pathogen interactions between different SARS-CoV-2 variants [26, 27], we performed univariate correlation analysis to test whether the causative viral variants disparately impacted immune responses to *A. fumigatus*. Background-corrected AfuLy-induced cytokine response was largely unaffected by the causative viral mutant (**Figure S7A-B**). However, there was a global trend toward more pronounced baseline hypercytokinemia in Delta patients (e.g., ρ > 0.8 for IL-4, IL-7, and IL-2R, **Figure S7A-B**). This trend also persisted in non-background-corrected AfuLy-reactive responses (**Figure S7A-B**).

For *A. fumigatus*-induced activation of antigen-presenting cells (APCs), we found a modest trend toward weaker reactogenicity to the fungus in Delta versus Alpha patients (e.g., ρ = -0.41 for ROS MFI, **Figure S7C**). This trend even reached significance when comparing individual background-adjusted frequencies of AfuG-reactive ROS- and CD62L-positive granulocytes (both p = 0.028, **Figure S7D**).

Furthermore, we tested the impact of 3 critical clinical parameters – maximum infection severity (WHO score), time since COVID-19 diagnosis (first positive PCR test), and GCS therapy – on anti-*Aspergillus* immunity. None of these parameters correlated significantly with key T-cellular parameters. However, GCS uptake showed modest positive correlation with PD-1 expression (ρ = 0.37, **Figure S8A**).

Maximum WHO scores correlated negatively with most baseline and AfuLy-reactive cytokine responses, whereas trends for background-adjusted AfuLy-induced responses were heterogenous (**Figure S8B**). GCS therapy expectedly counteracted baseline hypercytokinemia, whereas the impact of GCS on background-adjusted AfuLy-induced responses was inconsistent and insignificant (**Figure S8B**), suggesting that impaired cytokine responses to AfuLy is not an artifact of GCS therapy. Baseline concentrations of most T-cellular cytokines tended to slightly decrease over time after COVID-19 infection, further supporting the exhaustion hypothesis, whereas innate hypercytokinemia showed an inverse trend (**Figure S8B**). Most background-adjusted AfuLy-induced APC-derived cytokine responses worsened with increasing time after COVID-19 infection, whereas trends for T-cellular cytokines were inconsistent (**Figure S8B**). IL-2 stood out as the only response being significantly restored over time (ρ = 0.68, p = 0.017) and MIG/CXCL9 showed a similar trend (**Figure S8B-C**).

Expectedly, increasing COVID-19 severity had a modest negative effect on most surrogates of innate immune cell activation and maturation, most notably on CD83 upregulation (ρ = -0.53, **Figure S8D**). Aligning with trends for cytokine responses, innate immune cell activation and maturation mostly worsened with increasing time post COVID-19 infection (**Figure S8D**). Interestingly, most surrogates of innate immune cell responsiveness, especially percentages of granulocytes producing ROS in response to AfuG, showed modest to strong improvement with increasing GCS uptake (ρ = 0.72, p = 0.011, **Figure S8D-E**). Of note, Dectin-1 is known to be suppressed after DC activation by *A. fumigatus* antigens [28]; therefore, inverse correlation trends support the conclusions regarding innate immune cell activation for all 3 studied clinical parameters.

Taken together, these data suggest that the observed alterations in anti-*Aspergillus* immune responses are conserved across the heterogenous patient cohort. Immune dysfunction was largely unaffected by the studied clinical characteristics, except for greater impairment of granulocyte function in patients during the Delta wave and an association of GCS uptake with improved baseline hypercytokinemia and restored granulocyte dysfunction.

### Comparison of anti-Mucoralean immunity in COVID-19 patients and controls

Inspired by the emerging reports of CAM [2], we additionally tested anti-Mucoralean immunity in patients during the Delta wave. To that end, we selected *R. arrhizus*, the most common causative Mucorales species in patients with CAM [2]. As was the case for AfuLy stimulation, frequencies of background-corrected CD154^+^ RarLy-reactive Th cells (median 0.13% vs. 0.09%, **Figure S9A**) and their phenotypes (**Figure S9B**) did not differ significantly in COVID-19 patients and controls. Diminished CD69 induction and enrichment of proliferative (Ki-67^+^) Th cells among antigen-reactive T cells, seen before with AfuLy (**Figure 2C+G**), were confirmed for RarLy stimulation (**Figure S9A+E**). However, statistical significance was only reached for Ki-67 in the smaller RarLy-based dataset. RarLy-stimulated WB from COVID-19 patients revealed trends of increased Th-cell exhaustion compared to controls, with MMRs of 1.47, 1.33, 1.45, and 1.24 for PD-1^+^ Th, PD-1 MFI on Th, PD-1^+^ RarLy-reactive cells, and PD-1 MFI on RarLy-reactive cells, respectively (**Figure S9C-F**).

Twenty out of the 35 tested background-corrected RarLy-induced cytokine responses were significantly weaker in COVID-19 patients than in controls, including most of the studied T-cellular cytokines and a number of APC-derived cytokines (**Figure 5A**). PCA identified two distinct clusters formed by RarLy-induced cytokine responses in patient and control samples, with non-overlapping 95% confidence areas (**Figure 5B**). Although baseline hypercytokinemia in unstimulated samples contributed to diminished background-adjusted antigen-induced release, we identified a subset of 7 cytokines (IFN-γ, IL-2, IL-5, IL-13, IL-22, IP-10/CXCL10, MIG/CXCL9) with both significantly lower RarLy-reactive and background-adjusted RarLy-induced responses (**Figure 5C**, red asterisks in **Figure 5A**). IL-2 and IL-22 had MMRs of 0.14 and 0.03, respectively, for background-adjusted RarLy-induced release in samples from COVID-19 patients versus controls (**Figure 5A+D**). IFN-γ, IL-5, IL-13, IP-10/CXCL10, and MIG/CXCL9 had no detectable background-adjusted RarLy-induced release in most patient samples (median 0 pg/mL, **Figure 5A+D**).

**Figure 5.**
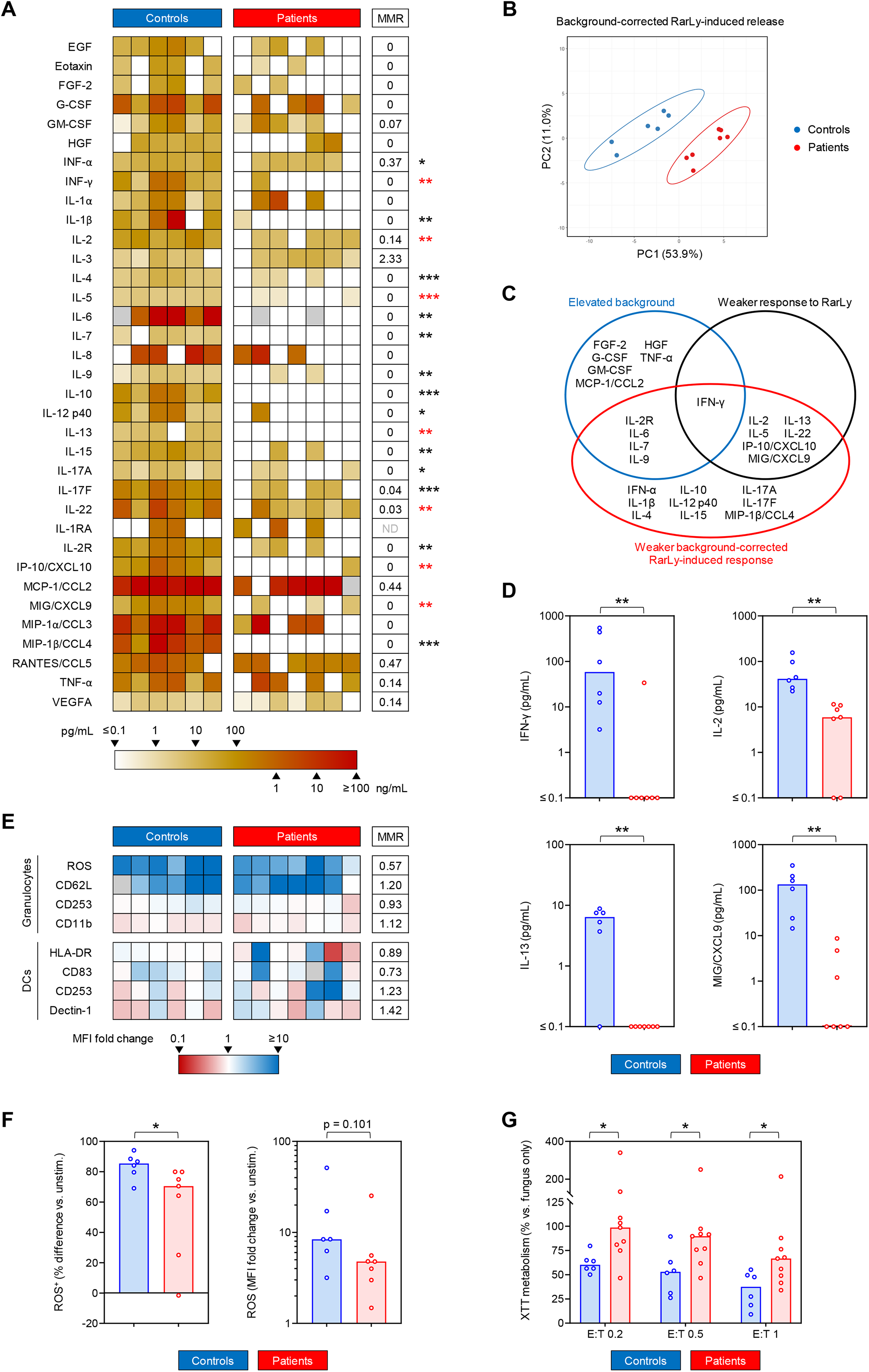
Patients with COVID-19 show impaired *Rhizopus arrhizus*-induced T-cellular cytokine response and weakened anti-*Rhizopus* activity of granulocytes. (**A**) Heat map representing individual background-adjusted *Rhizopus arrhizus* lysate (RarLy)-induced cytokine release. Grey boxes indicate non-determinable values (i.e., measurements with unstimulated background exceeding the detectable range). MMR = median-to-median ratio (patients/controls). ND = MMR not defined (median 0 pg/mL in both cohorts). (**B**) Principal component analysis (PCA) comparing background-adjusted RarLy-induced cytokine release in COVID-19 patients and controls. Ellipses represent 95% confidence ranges. (**C**) Venn diagram summarizing statistically significant alterations in unstimulated background, RarLy-reactive, and background-adjusted RarLy-induced cytokine responses in COVID-19 patients versus control. Significance was defined as p < 0.05 and false-positive discovery rate (FDR) < 0.2. (**D**) Individual and median (columns) background-adjusted RarLy-induced concentrations of selected cytokines. (**E**) Heat map summarizing induction of activation markers in/on granulocytes and dendritic cells (DCs) upon stimulation with *Rhizopus arrhizus* germlings (RarG). Fold changes of mean fluorescence intensity (MFI) in stimulated versus unstimulated samples are represented by color scale. Grey boxes indicate non-determinable ratios (i.e., measurements with a baseline MFI ≤ 0). (**F**) Individual and median (columns) differential frequencies of ROS-producing granulocytes and ROS MFI in RarG-stimulated whole blood (WB) versus unstimulated WB. (**G**) Relative XTT metabolism of *R. arrhizus* in the presence of neutrophils at different effector/target (E:T) ratios, normalized to a “fungus only” control without neutrophils. Lower values indicate stronger killing potential of neutrophils. (**A, D-G**), Mann-Whitney U test (patients versus controls) and Benjamini-Hochberg procedure to test for an FDR of < 0.2. * p < 0.05, ** p < 0.01, *** p < 0.001.

Expression intensity (MFI) of activation markers on granulocytes and DCs in response to RarG showed no significant differences between patients and controls based on the limited number of samples, although a trend toward weaker ROS production by granulocytes from COVID-19 patients was found (MMR 0.57, p = 0.101, **Figure 5E-F**). This trend was paralleled by a significantly lower number of granulocytes with a detectable ROS response to RarG in patients versus controls (70.5% vs. 85.4%, p = 0.035, **Figure 5F**). Furthermore, neutrophils isolated from COVID-19 patients had significantly weaker fungicidal activity against *R. arrhizus* than cells from control subjects, with MMRs of 1.56-1.63 for hyphal proliferation (**Figure 5G**, p = 0.029-0.043). Taken together, these findings indicate that samples from COVID-19 patients challenged with *R. arrhizus* antigens displayed severely impaired cytokine release, signals of Th-cell exhaustion, weakened neutrophilic ROS production, and less potent fungicidal activity against *R. arrhizus*.

### Pathway analysis and composite model of impaired anti-mold immunity in patients with COVID-19

In order to extrapolate an overall model of antifungal immune impairment in COVID-19 patients, we performed pathway enrichment analysis. As evident from the raw data, unsupervised enrichment analysis confirmed both exuberant baseline inflammation and increased T-cellular exhaustion in unstimulated samples of COVID-19 patients (**Figure 6A**). Furthermore, enrichment analysis confirmed broad impairment of all major background-adjusted Th responses to *A. fumigatus* and *R. arrhizus* antigens, with overall more diminished enrichment for responses to *R. arrhizus* (**Figure 6A**). Based on trends in cytokine responses and flow cytometric markers, enrichment analysis further predicted signals of weakened immune cell activation via PRR signaling and impaired crosstalk of various leukocyte populations (e.g., NK-cell/DC crosstalk) in mold-stimulated blood samples from COVID-19 patients (**Figure 6A**).

**Figure 6.**
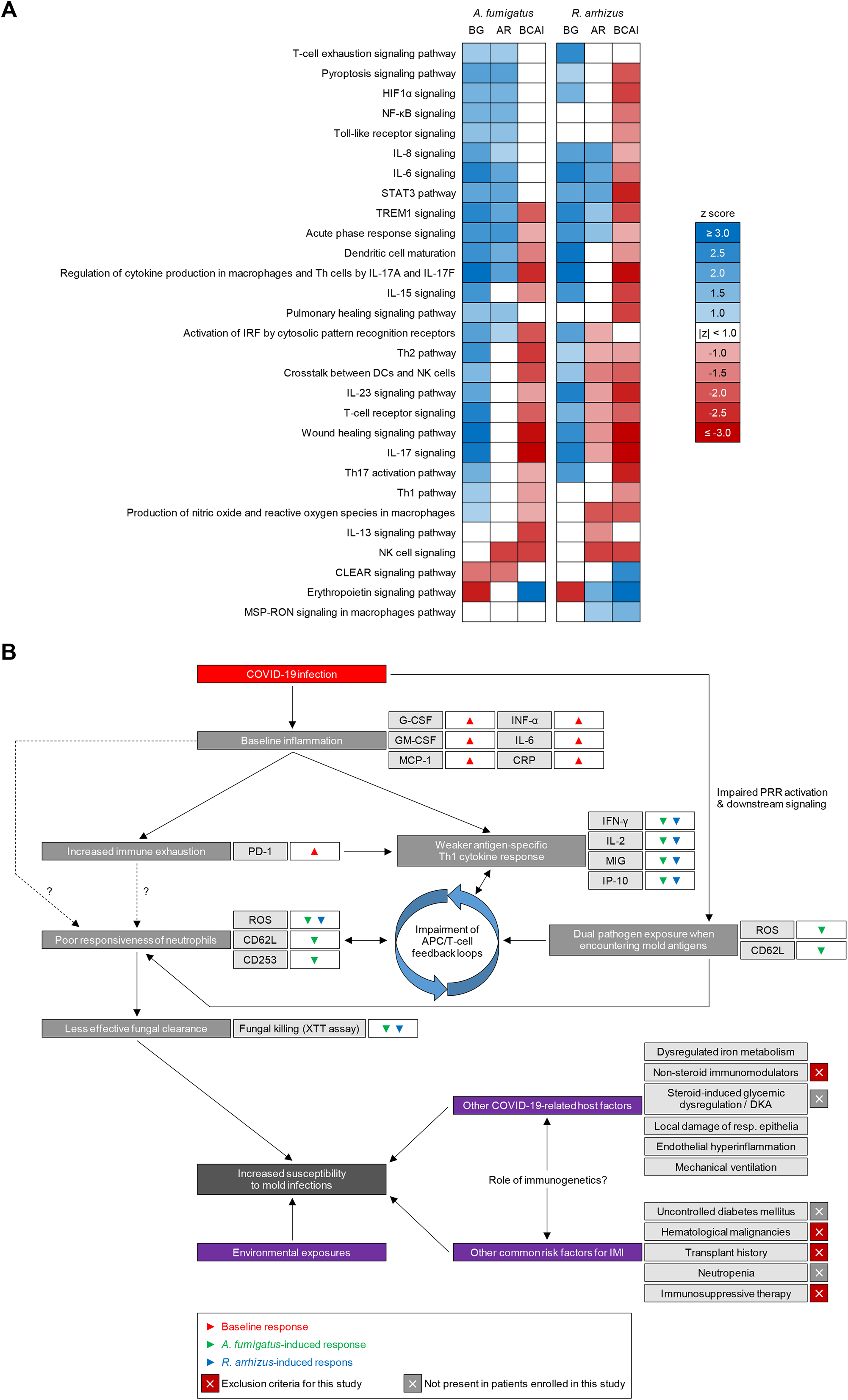
Patients with COVID-19 share common surrogates of impaired immune responses to opportunistic molds. (**A**) Activated (blue, z-score ≥ 1) and inhibited (red, z-score ≤ -1) cellular immune response and cytokine signaling pathways overrepresented by enrichment analysis of molecular response markers from multiplex cytokine assays and flow cytometry. Non-significant enrichments (absolute z-scores <1) are indicated in white. BG = background, AR = antigen-reactive, BCAI = background-corrected antigen-induced responses. (**B**) Schematic summarizing putative mechanisms of increased susceptibility to mold pathogens in patients with COVID-19. <u>Abbreviations:</u> HIF-1α = Hypoxia-Inducible Factor 1-alpha, NF-κB = Nuclear Factor kappa B, Th = T-helper cell(s), STAT3 = Signal Transducer and Activator of Transcription 3, TREM1 = Triggering Receptor Expressed on Myeloid cells 1, CLEAR = Coordinated Lysosomal Expression and Regulation, MSP-RON = Macrophage Stimulating Protein – Recepteur d’Origine Nantais, DKA = diabetic ketoacidosis.

Altogether, our results support a model whereby baseline hypercytokinemia, signals of immune exhaustion, reduced reactogenicity of innate immune cells (especially granulocytes), impaired cytokine responses to mold antigens, and potential disruption of intercellular feedback loops, contribute to less potent antifungal immunity in COVID-19 patients (**Figure 6B**). Potentially synergizing with other infection- and therapy-related effects on post-COVID-19 host defense and additional underlying risk factors, these severe immune alterations likely contribute to the predisposition of patients with and after COVID-19 to invasive mold infections (**Figure 6B**).

## Discussion

Although various risk factors for secondary fungal infections in COVID-19 patients have been described [2, 6, 10], direct experimental evidence for alterations of antifungal immunity has been lacking. Yet, identification of immunopathogenic links between the viral infection and predisposition to fungal co-infections will be essential to guide the development and use of immunomodulatory drugs in the treatment of post-viral mold infections. Herein, we identified three major mechanisms of diminished immune responses to two major mold pathogens, *A. fumigatus* and *R. arrhizus*, in COVID-19 patients: i) increased Th exhaustion with signs of reduced Th activation, ii) decreased mold-induced Th cytokine response, and iii) neutrophil dysfunction with declined fungicidal activity. The magnitude and strong conservation of the observed immune alterations in our heterogenous cohort with moderate disease severity suggests that impaired reactogenicity to molds is not only a consequence of a systemic stress response or exogenous factors (e.g., GCS therapy), but at least partly induced by a generic effect of the underlying SARS-CoV-2 infection.

SARS-CoV-2 promotes an imbalance of the cytokine milieu in the lung and bloodstream [29]. This hyperinflammatory state is driven by IL-6 and a variety of other cytokines/chemokines that showed strongly elevated baseline levels in our patient cohort. Local hyperinflammation in the pulmonary environment, sustained by neutrophils, monocytes, and macrophages, can lead to impaired integrity of the blood/air barrier and increased vascular permeability, resulting in alveolar edema and hypoxia [9, 30]. This is especially the case if antiviral Th-cell commitment, in combination with CTL activation and the B-cell-driven response, was not encompassing [31]. In this case, viral persistence and a consequent prolongation and amplification of innate immune mechanisms, associated with dysfunctional adaptive responses, can cause a ‘hyperinflammatory’ state underlying a so-called cytokine storm [32].

In addition to cytokine/chemokine-mediated lung injury, hyperinflammation can adversely shape the systemic immune environment [9]. For instance, it has been hypothesized that the susceptibility to fungal pathogens is partially linked to dysregulated interferon signaling [9]. Sustained production of high levels of type I interferons (i.e., IFN-α/β) can result in lymphopenia, weakened Th-cell activation, and immune exhaustion [33-35]. Our patient cohort had 2-fold higher baseline levels of IFN-α and elevations of IFN-α tended to negatively correlate with Th responses (data not shown), supporting a central role of overzealous type I interferon release in the immune pathogenesis of CAPA and CAM.

Dysregulated interferon signaling combined with increased baseline release of colony-stimulating factors and other proinflammatory cytokines and chemokines (e.g., RANTES/CCL5), as seen in our cohort, could provide a mechanistic link between altered cytokine signaling and the observed granulocyte dysfunction. Specifically, dysregulated myelopoiesis in COVID-19 patients can promote the release of increased numbers of immature and dysfunctional neutrophils [36]. In fact, blood counts and flow cytometric analyses in our patient cohort revealed both increased granulocyte production and a trend toward lower expression of the maturation marker CD16. Neutrophil dysfunction was likely further compounded by the observed diminished antigen-induced Th1 and Th17 responses. Inversely, the APC cytokine IP10/CXCL10, which was less potently secreted in response to mold antigens in our patient cohort, is a strong chemoattractant for T cells, monocytes, and NK cells [37]. Given the use of a WB-based assay instead of isolated cell populations for most assays shown in this manuscript, such feedback loops were captured by our approach [13] and enrichment analysis further corroborated signs of impaired crosstalk between key APC and Th populations (**Figure 6A**).

Importantly, our study specifically focused on direct post-COVID-19 impedance of mold antigen-reactive immunity in an *ex-vivo* setting. Therefore, our approach was not designed to investigate other, partially pre-described features of impaired antifungal defense in COVID-19 patients, such as pulmonary pathophysiology (discussed above), the role of endothelial inflammation, or the systemic *in-vivo* impact of iatrogenic interventions. Consequently, our findings only cover a small yet potentially impactful portion of the complex co-pathogenesis of COVID-19 and opportunistic mold infections (**Figure 6B**).

GCS are currently recommended by U.S. and European treatment guidelines to minimize systemic inflammation and mitigate overzealous lung tissue damage in COVID-19 patients requiring supplemental oxygen [38]. Our observation that GCS tended to counteract deficient ROS response of granulocytes aligns with the results of a recent study showing that steroid-mediated attenuation of the hyperinflammatory environment in COVID-19 patients improved the myeloid cell effector response against bacterial challenge [39]. However, the overwhelming clinical evidence that GCS increase the risk of CAPA and especially CAM [1, 2, 6, 10] clearly indicates that the protective impact of GCS on individual aspects of mold-reactive immune responses is strongly outweighed by adverse systemic effects of GCS on host metabolism, immune defense, and expression of epithelial receptors promoting fungal invasion [2, 10, 40, 41]. Additionally, we found modest positive correlation of steroid uptake with PD-1 expression on Th cells, which aligns with the well-known role of GCS as a driver of immune exhaustion and checkpoint pathway expression [42]. Taken together, our data should not be interpreted as suggestive of a protective effect of GCS but rather as an indication of additional, largely steroid-independent signs of impaired antifungal immunity due to the underlying COVID-19 infection.

Our findings of steroid-independent alterations of mold antigen-specific immunity would also encourage further comparative studies in other post-viral infection settings (e.g., after influenza pneumonia). We currently have a very limited understanding how i) general stress responses due to severe infection and illness, ii) shared motifs of post-viral immune alterations, iii) infection-specific immune features, iv) iatrogenic interventions, and v) underlying host predisposition differentially intersect in the nuanced co-pathogenesis of CAPA, CAM, and other entities such as IAPA. As extensively reviewed elsewhere [9, 43], there are key differences in the immune pathogenesis of post-viral mycoses depending on the underlying viral infection. For instance, influenza tends to cause more severe epithelial lysis than COVID-19, contributing to greater predisposition to fungal angioinvasion [44]. Furthermore, onset of CAPA and CAM often has a longer lag-time after the underlying viral pneumonia than IAPA [3], potentially suggesting a relatively more profound role of immune exhaustion in post-COVID-19 mycoses than in IAPA. Additionally, global differences between these entities are likely considerably modulated by strain-specific differences in the viral interaction with host immunity, as suggested by the influenza literature [45] and our comparison of antifungal immune features in patients infected with the Alpha versus Delta SARS-CoV-2 variants.

Due to the restrictive exclusion criteria, a major limitation of this study was the relatively small cohort size, especially for responses to *R. arrhizus* that were only tested during the Delta wave (after the emergence of CAM). The high consistency of our findings, among a heterogenous patient cohort, reduces the impact of this limitation on conclusions regarding global alterations of anti-mold immunity in COVID-19 patients, but downstream comparisons of immune impairment with specific clinical characteristics were severely underpowered and mostly relied on “soft statistics” such as correlation. This limitation is further compounded by the fact that some of the clinical variables were not entirely independent. For instance, due to the restriction of our study to patients with moderate disease severity (WHO scores 4 and 5) at the time of immune cell sampling, patients with higher initial COVID-19 severity tended to be enrolled at a later stage of their post-COVID-19 recovery (after release from the ICU). The size of our dataset precluded multivariate analysis to determine the independent impact of these and other clinical characteristics on immune readouts. Furthermore, the limited available blood volume precluded both a detailed wet-lab confirmation of the underlying immune pathways predicted by enrichment analysis and the assessment of immune responses to additional (i.e., non-fungal) antigens. Therefore, some of the observed immune alterations are likely not specific to fungal antigens, as suggested by published evidence regarding impairment of granulocyte responses to *Staphylococcus aureus* and *Streptococcus pneumoniae* in COVID-19 patients [39, 46]. Lastly, immunogenetic factors could have contributed to highly variable CAPA incidence rates and the regional emergence of CAM in South Asia [47]. Such factors cannot be sufficiently captured by our small study with a local catchment area; thus, our results would need to be validated in well-controlled multi-center studies involving patients from different geographical areas and immunogenetic backgrounds.

Despite these limitations, this study provides an inaugural characterization of impaired immune responses to *A. fumigatus* and *R. arrhizus* antigens in patients with and after COVID-19 pneumonia, beyond the well-studied adverse impact of GCS. The predicted immune pathways will serve as a hypothesis-generating foundation for detailed follow-up studies of the immune predisposition to secondary mold infections after respiratory viral infections, including COVID-19, influenza, as well as infections due to respiratory syncytial virus, parainfluenza, and adenovirus. If further validated in multi-center studies and/or suitable animal models, the identified pathways can open new avenues for immunotherapeutic strategies to prevent and treat fungal superinfections in patients with post-viral immune impairment.

## Data Availability

All data produced in the present study are available upon reasonable request to the authors.

## Acknowledgements

The authors wish to thank all blood donors.

This work was supported by a grant of the “Stiftung zur Förderung der Krebsforschung an der Universität Würzburg (“Forschung hilft”)”, grant allocation number 8607630 (to CDL and JL); "Nachweis von Immunzellbotenstoffen gegen Covid-19 für die Diagnostik und Impfstofftestung”. The study was further supported by the Deutsche Forschungsgemeinschaft (DFG) within the Collaborative Research Center CRC124 FungiNet “Pathogenic fungi and their human host: Networks of interaction,” DFG project number 210879364 (project A1 to AAB, A2 to HE and JL, C3 to OKu, and Z2 to OKn).

## Author Contributions

The study was conceived by BT, CDL, JL, and SW. Patient enrollment and clinical documentation were performed by SK, EPJ, LB, LP, and MH. Experiments were planed and performed by BT, LS, SR, and KH. Resources were provided by OKn and AAB. Data were analyzed by JPG, SS, and SW. Data were visualized by SW. Project administration and supervision were led by CDL, OKu, GP, LPW, HE, JL, and SW. Funding was acquired by CDL, HE, and JL. The original draft was written by BT, CDL, JL, and SW. All co-authors reviewed, edited, and approved the manuscript.

## References

1. Arastehfar, A., et al., COVID-19 Associated Pulmonary Aspergillosis (CAPA)-From Immunology to Treatment. J Fungi (Basel), 2020. 6(2).

2. Hoenigl, M., et al., The emergence of COVID-19 associated mucormycosis: a review of cases from 18 countries. The Lancet. Microbe, 2022.

3. Feys, S., et al., A Visual and Comprehensive Review on COVID-19-Associated Pulmonary Aspergillosis (CAPA). Journal of Fungi, 2021. 7(12).

4. Russell, C.D., J.E. Millar, and J.K. Baillie, Clinical evidence does not support corticosteroid treatment for 2019-nCoV lung injury. Lancet, 2020. 395(10223): p. 473–475.

5. Cox, M.J., et al., Co-infections: potentially lethal and unexplored in COVID-19. Lancet Microbe, 2020. 1(1): p. e11.

6. Koehler, P., et al., COVID-19 associated pulmonary aspergillosis. Mycoses, 2020. 63(6): p. 528–534.

7. Alanio, A., et al., Prevalence of putative invasive pulmonary aspergillosis in critically ill patients with COVID-19. Lancet Respir Med, 2020. 8(6): p. e48–e49.

8. Rutsaert, L., et al., COVID-19-associated invasive pulmonary aspergillosis. Annals of Intensive Care, 2020. 10(1): p. 71.

9. Salazar, F., et al., Pathogenesis of Respiratory Viral and Fungal Coinfections. Clin Microbiol Rev, 2022. 35(1): p. e0009421.

10. Muthu, V., et al., Epidemiology and Pathophysiology of COVID-19-Associated Mucormycosis: India Versus the Rest of the World. Mycopathologia, 2021. 186(6): p. 739–754.

11. Claypool, K.T., et al., Characteristics of undiagnosed diabetes in men and women under the age of 50 years in the Indian subcontinent: the National Family Health Survey (NFHS-4)/Demographic Health Survey 2015–2016. BMJ Open Diabetes Research &amp;amp; Care, 2020. 8(1): p. e000965.

12. A minimal common outcome measure set for COVID-19 clinical research. Lancet Infect Dis, 2020. 20(8): p. e192–e197.

13. Lauruschkat, C.D., et al., Development of a Simple and Robust Whole Blood Assay with Dual Co-Stimulation to Quantify the Release of T-Cellular Signature Cytokines in Response to Aspergillus fumigatus Antigens. J Fungi (Basel), 2021. 7(6).

14. Hünniger, K., et al., A Virtual Infection Model Quantifies Innate Effector Mechanisms and Candida albicans Immune Escape in Human Blood. PLOS Computational Biology, 2014. 10(2): p. e1003479.

15. Metsalu, T. and J. Vilo, ClustVis: a web tool for visualizing clustering of multivariate data using Principal Component Analysis and heatmap. Nucleic Acids Research, 2015. 43(W1): p. W566–W570.

16. Krämer, A., et al., Causal analysis approaches in Ingenuity Pathway Analysis. Bioinformatics, 2014. 30(4): p. 523–30.

17. Arens, R. and S.P. Schoenberger, Plasticity in programming of effector and memory CD8+ T-cell formation. Immunological Reviews, 2010. 235(1): p. 190–205.

18. Jameson, S.C. and D. Masopust, Diversity in T Cell Memory: An Embarrassment of Riches. Immunity, 2009. 31(6): p. 859–871.

19. Weis, P., et al., Development and evaluation of a whole blood-based approach for flow cytometric quantification of CD154+ mould-reactive T cells. Med Mycol, 2020. 58(2): p. 187–196.

20. Bacher, P., et al., Human Anti-fungal Th17 Immunity and Pathology Rely on Cross-Reactivity against Candida albicans. Cell, 2019. 176(6): p. 1340-1355.e15.

21. Page, L., et al., Evaluation of Aspergillus and Mucorales specific T-cells and peripheral blood mononuclear cell cytokine signatures as biomarkers of environmental mold exposure. Int J Med Microbiol, 2018. 308(8): p. 1018–1026.

22. Lauruschkat, C.D., et al., Chronic Occupational Mold Exposure Drives Expansion of Aspergillus-Reactive Type 1 and Type 2 T-Helper Cell Responses. J Fungi (Basel), 2021. 7(9).

23. Wurster, S., et al., Intra-and inter-individual variability of Aspergillus fumigatus reactive T-cell frequencies in healthy volunteers in dependency of mould exposure in residential and working environment. Mycoses, 2017. 60(10): p. 668–675.

24. Stephen-Victor, E., et al., Aspergillus fumigatus Cell Wall α-(1,3)-Glucan Stimulates Regulatory T-Cell Polarization by Inducing PD-L1 Expression on Human Dendritic Cells. J Infect Dis, 2017. 216(10): p. 1281–1294.

25. Tobin, J.M., et al., Influenza Suppresses Neutrophil Recruitment to the Lung and Exacerbates Secondary Invasive Pulmonary Aspergillosis. J Immunol, 2020. 205(2): p. 480–488.

26. McCallum, M., et al., Molecular basis of immune evasion by the Delta and Kappa SARS-CoV-2 variants. Science, 2021. 374(6575): p. 1621–1626.

27. Bojkova, D., et al., Reduced interferon antagonism but similar drug sensitivity in Omicron variant compared to Delta variant of SARS-CoV-2 isolates. Cell Research, 2022. 32(3): p. 319–321.

28. Hellmann, A.-M., et al., Human and Murine Innate Immune Cell Populations Display Common and Distinct Response Patterns during Their In Vitro Interaction with the Pathogenic Mold Aspergillus fumigatus. Frontiers in Immunology, 2017. 8.

29. Rabaan, A.A., et al., Role of Inflammatory Cytokines in COVID-19 Patients: A Review on Molecular Mechanisms, Immune Functions, Immunopathology and Immunomodulatory Drugs to Counter Cytokine Storm. Vaccines (Basel), 2021. 9(5).

30. Channappanavar, R. and S. Perlman, Pathogenic human coronavirus infections: causes and consequences of cytokine storm and immunopathology. Semin Immunopathol, 2017. 39(5): p. 529–539.

31. Ahmadpoor, P. and L. Rostaing, Why the immune system fails to mount an adaptive immune response to a COVID-19 infection. Transpl Int, 2020. 33(7): p. 824–825.

32. Sarzi-Puttini, P., et al., COVID-19, cytokines and immunosuppression: what can we learn from severe acute respiratory syndrome? Clin Exp Rheumatol, 2020. 38(2): p. 337–342.

33. Shiow, L.R., et al., CD69 acts downstream of interferon-alpha/beta to inhibit S1P1 and lymphocyte egress from lymphoid organs. Nature, 2006. 440(7083): p. 540–4.

34. Sumida, T.S., et al., Type I interferon transcriptional network regulates expression of coinhibitory receptors in human T cells. Nature Immunology, 2022. 23(4): p. 632–642.

35. McNab, F., et al., Type I interferons in infectious disease. Nature Reviews Immunology, 2015. 15(2): p. 87–103.

36. Schulte-Schrepping, J., et al., Severe COVID-19 Is Marked by a Dysregulated Myeloid Cell Compartment. Cell, 2020. 182(6): p. 1419-1440.e23.

37. Pelaia, C., et al., Lung under attack by COVID-19-induced cytokine storm: pathogenic mechanisms and therapeutic implications. Ther Adv Respir Dis, 2020. 14: p. 1753466620933508.

38. Health, N.I.o. Figure 2. Therapeutic Management of Adults Hospitalized for COVID-19 Based on Disease Severity. 2022. 04/08/2022; 11.43 AM; Available from: https://www.covid19treatmentguidelines.nih.gov/management/clinical-management/hospitalized-adults--therapeutic-management/hospitalized-adults-figure/.

39. Mairpady Shambat, S., et al., Hyperinflammatory environment drives dysfunctional myeloid cell effector response to bacterial challenge in COVID-19. PLOS Pathogens, 2022. 18(1): p. e1010176.

40. Leistner, R., et al., Corticosteroids as risk factor for COVID-19-associated pulmonary aspergillosis in intensive care patients. Crit Care, 2022. 26(1): p. 30.

41. Bentvelsen, R.G., et al., Regional Impact of COVID-19-Associated Pulmonary Aspergillosis (CAPA) during the First Wave. J Fungi (Basel), 2022. 8(2).

42. Adorisio, S., et al., Glucocorticoid and PD-1 Cross-Talk: Does the Immune System Become Confused? Cells, 2021. 10(9).

43. Lamoth, F., et al., Navigating the uncertainties of COVID-19 associated aspergillosis (CAPA): A comparison with influenza associated aspergillosis (IAPA). J Infect Dis, 2021.

44. van de Veerdonk, F.L., et al., COVID-19-associated Aspergillus tracheobronchitis: the interplay between viral tropism, host defence, and fungal invasion. The Lancet Respiratory Medicine, 2021. 9(7): p. 795–802.

45. Chen, X., et al., Host Immune Response to Influenza A Virus Infection. Front Immunol, 2018. 9: p. 320.

46. Nomani, M., et al., Decreased neutrophillmediated bacterial killing in COVIDl19 patients. Scandinavian Journal of Immunology, 2021: p. e13083.

47. Chakrabarti, S.S., et al., The Pathogenetic Dilemma of Post-COVID-19 Mucormycosis in India. Aging Dis, 2022. 13(1): p. 24–28.

