## Supplementary Figure Legends for "COVID-19 patients share common, corticosteroid-independent features of impaired host immunity to pathogenic molds"

#### Supplementary Figure Legends and Tables


**Authors:**

Beeke Tappe[1,*] and Chris D. Lauruschkat[1,*], Lea Strobel[1], Jezreel Pantaleón García[2], Oliver Kurzai[3,4], Silke Rebhan[1], Sabrina Kraus[1], Elena Pfeuffer-Jovic[5], Lydia Bussemer[1], Lotte Possler[6], Matthias Held[5], Kerstin Hünniger[3,4], Olaf Kniemeyer[4], Sascha Schäuble[4], Axel A. Brakhage[4,7], Gianni Panagiotou[4], P. Lewis White[8], Hermann Einsele[1], Jürgen Löffler[1, $,#] and Sebastian Wurster[9,#]

**Affiliations:**

1 Department of Internal Medicine II, University Hospital of Würzburg, Würzburg, Germany.

2 Department of Pulmonary Medicine, The University of Texas MD Anderson Cancer Center, Houston, USA.

3 Institute for Hygiene and Microbiology, University of Würzburg, Würzburg, Germany.

4 Leibniz Institute for Natural Product Research and Infection Biology–Hans Knöll Institute, Jena, Germany.

5 Missionsärztliche Klinik Würzburg, Würzburg, Germany.

6 Main-Klinik Ochsenfurt, Würzburg, Germany.

7 Department of Microbiology and Molecular Biology, Institute of Microbiology, Friedrich Schiller University, Jena, Germany

8 Public Health Wales, Microbiology Cardiff, Wales, UK.

9 Department of Infectious Diseases, Infection Control and Employee Health, The University of Texas MD Anderson Cancer Center, Houston, USA.

* Shared first authors

$ Requests for materials should be addressed to Jürgen Löffler

### Shared last authors and co-corresponding authors:

**Prof. Dr. Jürgen Löffler**
Department of Internal Medicine II
University Hospital of Würzburg
Josef Schneider Str. 2, C11, 97080 Würzburg, Germany


**Asst. Prof. Dr. Sebastian Wurster**
Department of Infectious Diseases, Infection Control and Employee Health
The University of Texas MD Anderson Cancer Center
1515 Holcombe Blvd, FCT 12.6048, Houston, TX 77030, USA


#### Supplementary Figure Legends

**Supplementary Figure 1. Gating strategy to study global and mold antigen-reactive T-cell responses (panel 1).**

Single cells were gated based on SSC-A, FSC-A, and FSC-H properties. Lymphocytes were identified by light scatter properties and dead lymphocytes were excluded by Live/Dead staining. $CD4^{+}$, $CD8^{bright}$, and $CD8^{dim}$ cells were differentiated. Within each T-cell subpopulation, positive cells for activation markers CD154 and CD69, exhaustion marker PD-1, and proliferation marker Ki-67 were gated. $CD4^{+}$ T-helper (Th) cells were subdivided into Th1 ($CXCR3^{+}$), Th2 ($CXCR3^{-}$ $CCR6^{-}$ $CCR4^{+}$), and Th17 ($CXCR3^{-}$ $CCR6^{+}$ $CCR4^{+}$) cells. Th-cell polarization was also determined for $CD154^{+}$, $CD69^{+}$, PD-$1^{+}$ and Ki-$67^{+}$ Th cells. Mean fluorescence intensity was determined for all markers.

**Supplementary Figure 2. Gating strategy to study global and mold antigen-reactive T-cell responses (panel 2).**

Single cells were gated based on SSC-A, FSC-A, and FSC-H properties. Lymphocytes were identified by light scatter properties and dead lymphocytes were excluded by Live/Dead staining. $CD4^{+}$, $CD8^{bright}$, and $CD8^{dim}$ cells were differentiated. Within each T-cell subpopulation, positive cells for activation markers CD154, CD69, and CD107a, exhaustion marker PD-1, and proliferation marker Ki-67 were gated. Effector memory T cells ($T_{EM}$, $CD45RA^{-}$ $CCR7^{-}$), central memory T cells ($T_{CM}$, $CD45RA^{-}$ $CCR7^{+}$), effector T cells ($T_{EMRA}$, $CD45RA^{+}$ $CCR7^{-}$) and naïve T cells ($T_{N}$, $CD45RA^{+}$ $CCR7^{+}$) were differentiated. Mean fluorescence intensity was determined for all markers.

**Supplementary Figure 3. Gating strategy to determine baseline leukocyte distributions and to study mold antigen-induced activation of dendritic cells and granulocytes.**

Debris was excluded by light scatter properties and dead cells were excluded by Live/Dead staining. For determination of leukocyte distributions (performed on unstimulated samples), T cells ($CD19^{-}$ $CD3^{+}$) and B cells ($CD19^{+}$ $CD3^{-}$) were gated. $CD19^{-}$ $CD3^{-}$ cells were further subdivided into monocytes ($CD14^{+}$) and granulocytes ($CD66b^{+}$). Natural killer (NK) cells were identified as $CD3^{-}$ $CD56^{+}$ and subdivided into CD56$^{dim}$ CD16$^{+}$ NK cells and CD56$^{bright}$ CD16$^{low}$ NK cells. Granulocytes were identified by CD66 expression and analyzed for ROS, CD253, CD16, CD11b, and CD62L expression. Dendritic cells, identified as CD1c$^{+}$ cells, were analyzed for Dectin-1, CD253, HLA-DR, CD83, CD284 and CD14 expression. Mean fluorescence intensity was determined for all markers.

**Supplementary Figure 4. Patients with COVID-19 exhibit strong hypercytokinemia in plasma supernatants of unstimulated whole blood.**

Heat map summarizing individual cytokine concentrations in unstimulated whole blood samples. MMR = median-to-median ratio (patients/controls). ∞ = infinite MMR (median 0 pg/mL in the control cohort). ▲ = high (>1000) yet undefined MMR (median of the patient cohort exceeding the quantifiable range). Mann-Whitney U test (patients versus controls) and Benjamini-Hochberg procedure to test for an FDR of $< 0.2$. * $p < 0.05$, ** $p < 0.01$.

**Supplementary Figure 5. Dual antigen exposure of whole blood from healthy donors cannot recapitulate impaired *Aspergillus*-induced cytokine response in COVID-19 patients.**

Heat map summarizing individual background-adjusted cytokine release in whole blood samples from healthy donor stimulated with *Aspergillus fumigatus* lysate (AfuLy) or a combination of AfuLy and SARS-CoV-2 Protein S (PrS). Grey boxes indicate non-determinable values (i.e., measurements with unstimulated background exceeding the detectable range). Paired Wilcoxon test and Benjamini-Hochberg procedure to test for a false-positive discovery rate (FDR) of $< 0.2$. No comparison reached FDR-adjusted significance. ▪ $p < 0.05$ and FDR $> 0.2$. Colors of squares indicate the condition eliciting stronger cytokine release.

**Supplementary Figure 6. Granulocytes and dendritic cells (DCs) from COVID-19 patients display a less mature phenotype.**

Individual and median (columns) expression (mean fluorescence intensity, MFI) of maturation markers and pattern recognition receptors on granulocytes (**A**) and DCs (**B**) in unstimulated whole blood. Mann-

Whitney U test (patients versus controls) and Benjamini-Hochberg procedure to test for an FDR of < 0.2. * $p < 0.05$.

**Supplementary Figure 7. Patients infected with the SARS-CoV-2 Delta variant have stronger baseline cytokinemia and weaker granulocyte responses to *Aspergillus fumigatus*.**

(**A**) Heat map summarizing background-adjusted *Aspergillus fumigatus* lysate (AfuLy)-induced cytokine release in COVID-19 patients during the Alpha and Delta wave. Additionally, rank-biserial correlation coefficients comparing unstimulated background (BG), AfuLy-reactive (AR), and background-corrected AfuLy-induced (BCAI) cytokine release with the wave (Alpha = 1, Delta = 2) are provided. Negative (green) and positive (orange) values indicate stronger cytokine responses in patients infected with the Alpha and Delta variant, respectively. Significant correlation is indicated by bold black fonts. Bold white fonts indicate correlation coefficients that remained significant after Benjamini-Hochberg correction for an FDR of 0.2. (**B**) Principal component analysis comparing unstimulated background, AfuLy-reactive, and background-adjusted AfuLy-induced cytokine release in patients during the Alpha and Delta wave. Ellipses represent 95% confidence ranges. (**C**) Heat map summarizing induction of activation markers in/on granulocytes and dendritic cells upon stimulation with *Aspergillus fumigatus* germlimgs (AfuG) in COVID-19 patients during the Alpha and Delta wave. Fold changes of mean fluorescence intensity (MFI) in stimulated versus unstimulated samples are represented by color scale. Grey boxes indicate non-determinable ratios (i.e., measurements with a baseline MFI ≤ 0). Rank-biserial correlation was performed as described above. (**D**) Background-adjusted AfuG-responsive frequencies of ROS- and CD62L-positive granulocytes in COVID-19 patients during the Alpha and Delta wave. Mann-Whitney U test. * $p < 0.05$.

**Supplementary Figure 8. Altered immune responses to *Aspergillus fumigatus* in hospitalized COVID-19 patients are largely decoupled from infection severity and glucocorticosteroid therapy.**

(**A**) Spearman rank correlation coefficients for comparisons of activation and exhaustion marker-positive frequencies of T-helper (Th) cells in *Aspergillus fumigatus* lysate (AfuLy)-stimulated whole blood with the patients' maximum WHO progression scores ("WHO"), glucocorticosteroid uptake within the past 7 days ("GCS", measured in mg total prednisolone equivalent), and time between first positive SARS-CoV-2 polymerase chain reaction (PCR) test and blood sampling for immunoassays ("Time"). CD154 and CD69 responses were background-adjusted. Asterisks in the first column indicate significantly weaker responses in COVID-19 patients ("P") versus controls ("C"), as shown in **Figure 2**. * $p < 0.05$, ** $p < 0.01$, *** $p < 0.001$. (**B**) Spearman rank correlation coefficients for comparisons of clinical characteristics with unstimulated background (BG), AfuLy-reactive (AR), and background-corrected AfuLy-induced (BCAI) cytokine release. Significant correlation is indicated by bold black fonts. Bold white fonts indicate correlation coefficients that remained significant after Benjamini-Hochberg correction for an FDR of 0.2. Asterisks indicate significantly weaker AfuLy-induced cytokine release in COVID-19 patients versus controls, as shown in **Figure 3A**. Red asterisks denote significantly weaker AfuLy-reactive and background-corrected AfuLy-induced responses in COVID-19 patients, additionally highlighted by red boxes. (**C**) Correlation plots comparing time after positive PCR and background-corrected AfuLy-induced IL-2 and MIG/CXCL9 release. Spearman rank correlation coefficients (ρ) and their p-values are provided. (**D**) Spearman rank correlation coefficients for comparisons of clinical characteristics with background-adjusted *Aspergillus fumigatus* germling (AfuG)-responsive frequencies of activation marker-positive granulocytes and fold changes of median fluorescence intensity (MFI) of activation markers on granulocytes and dendritic cells (DCs) in AfuG-stimulated versus unstimulated whole blood. Asterisks indicate significantly weaker responses in COVID-19 patients versus controls, as shown in **Figures 4A**. (**E**) Correlation plots comparing steroid intake within the past 7 days and background-adjusted ROS response to AfuG. Spearman rank correlation coefficients (ρ) and their p-values are given.

**Supplementary Figure 9. COVID-19 patients show signs of impaired *Rhizopus arrhizus*-induced T-cell activation and increased T-helper cell exhaustion.**

(**A**) Background-corrected frequencies of *Rhizopus arrhizus* lysate (RarLy)-reactive $CD4^+$ T-helper (Th) cells detectable by CD154 or CD69 upregulation. (**B**) Mean distributions of memory/effector phenotypes and polarization of RarLy-reactive Th cells. (**C**) Frequencies of PD-$1^+$ cells among Th cells in RarLy-stimulated samples. (**D**) Mean fluorescence intensity (MFI) of PD-1 on Th cells in RarLy-stimulated samples. (**E**) Frequencies of $CD69^+$, $Ki\text{-}67^+$, and $PD\text{-}1^+$ cells among $CD4^+$ $CD154^+$ RarLy-reactive Th cells. (**F**) PD-1 MFI of $CD4^+$ $CD154^+$ RarLy-reactive Th cells. (**A, C-F**) Columns represent medians. (**A-F**) Mann-Whitney U test (patients versus controls) and Benjamini-Hochberg procedure to test for a false-positive discovery rate (FDR) of < 0.2. ** $p < 0.01$. Abbreviations: $T_{CM}$ = central memory T cells, $T_{EM}$ = effector memory T cells, $T_{EFF}/T_{EMRA}$ = effector T cells/terminally differentiated effector memory T cells re-expressing CD45RA.

**Supplementary Table 1. Preparation of whole blood stimulation tubes for *ex-vivo* immunoassays.**

| | **Assay** | **α-CD28** | **α-CD49d** | **AfuLy** | **AfuG** | **RarLy** | **RarG** | **PrS** | **PHA** | **RPMI** |
|---|---|---|---|---|---|---|---|---|---|---|
| **Concentration in ready-to-use stimulation tubes** | n/a | 2 µg/mL | 2 µg/mL | 100 µg/mL | $1{\times}10^{6}$ AfuG/mL | 100 µg/mL | $1{\times}10^{6}$ RarG/mL | 1.2 nmol | 10 µg/mL | *Ad* 500 µL |
| **Final concentration after injection of 500 µL WB** | n/a | 1 µg/mL | 1 µg/mL | 50 µg/mL | $5{\times}10^{5}$ AfuG/mL | 50 µg/mL | $5{\times}10^{5}$ RarG/mL | 0.6 nmol | 5 µg/mL | n/a |
| **Unstimulated control** | AI, Cyt | X | X | | | | | | | X |
| | II | | | | | | | | | X |
| **AfuLy stimulation** | AI, Cyt | X | X | X | | | | | | X |
| **AfuG stimulation** | II | | | | X | | | | | X |
| **RarLy stimulation** | AI, Cyt | X | X | | | X | | | | X |
| **RarG stimulation** | II | | | | | | X | | | X |
| **PrS stimulation** | AI, Cyt | X | X | | | | | X | | X |
| | II | | | | | | | X | | X |
| **AfuLy + PrS stimulation** | AI, Cyt | X | X | X | | | | X | | X |
| **AfuG + PrS stimulation** | II | | | | X | | | X | | X |
| **Positive control** | AI, II, Cyt | | | | | | | | X | X |

X indicates that the compound was used for the respective condition.

Abbreviations: AfuLy = *A. fumigatus* mycelial lysate, RarLy = *R. arrhizus* mycelial lysate, CD = cluster of differentiation, n/a = not applicable, PrS = SARS-CoV-2 Protein S (Alpha variant), PHA = phytohemagglutinin, RPMI = Roswell Park Memorial Institute medium, WB = whole blood, AI = adaptive immunity, II = innate immunity, Cyt = multiplex cytokine assay.

**Supplementary Table 2. Antibodies used for flow cytometric analyses.**

| Name | Fluorochrome | Clone | Provider | Catalogue number | Panel(s) |
|---|---|---|---|---|---|
| **CD1c (BDCA-1)** | PerCP-Vio700 | REA694 | Miltenyi Biotec | 130-110-538 | DC |
| **CD3** | PE-Vio615 | REA613 | Miltenyi Biotec | 130-114-520 | ID |
| **CD4** | VioGreen | REA623 | Miltenyi Biotec | 130-113-230 | T1, T2 |
| **CD8** | PerCP-Vio700 | REA734 | Miltenyi Biotec | 130-110-682 | T1, T2 |
| **CD11b** | APC | REA713 | Miltenyi Biotec | 130-110-554 | GR |
| **CD14** | APC-Vio770 | TÜK4 | Miltenyi Biotec | 130-113-144 | ID, DC |
| **CD16** | VioGreen | REA423 | Miltenyi Biotec | 130-113-397 | ID, GR |
| **CD19** | PE | REA675 | Miltenyi Biotec | 130-113-646 | ID |
| **CD45RA** | PE-Vio615 | REA1047 | Miltenyi Biotec | 130-117-745 | T2 |
| **CD56** | FITC | REA196 | Miltenyi Biotec | 130-114-549 | ID |
| **CD62L** | PE | REA615 | Miltenyi Biotec | 130-113-625 | GR |
| **CD66b** | PerCP-Vio700 | REA306 | Miltenyi Biotec | 130-119-768 | ID, GR |
| **CD69** | APC-Vio770 | REA824 | Miltenyi Biotec | 130-112-616 | T1, T2 |
| **CD83** | FITC/ VioBright515 | REA714 | Miltenyi Biotec | 130-110-507 | DC |
| **CD107a (LAMp-1)** | APC/R667 | REA792 | Miltenyi Biotec | 130-111-626 | T2 |
| **CD154** | FITC/ VioBright515 | REA238 | Miltenyi Biotec | 130-122-800 | T1, T2 |
| **CD183 (CXCR3)** | APC | REA232 | Miltenyi Biotec | 130-120-450 | T1 |
| **CD194 (CCR4)** | PE | REA279 | Miltenyi Biotec | 130-120-456 | T1 |
| **CD196 (CCR6)** | PE-Vio615 | REA190 | Miltenyi Biotec | 130-120-459 | T1 |
| **CD197 (CCR7)** | PE | REA546 | Miltenyi Biotec | 130-119-583 | T2 |
| **CD253 (TRAIL)** | PE-Vio615 | REA1113 | Miltenyi Biotec | 130-119-286 | GR, DC |
| **CD279 (PD-1)** | PE-Vio770 | REA1165 | Miltenyi Biotec | 130-120-385 | T1, T2 |
| **CD284 (TLR4)** | APC | HTA125 | Miltenyi Biotec | 130-096-236 | DC |
| **Dectin-1** | PE | REA515 | Miltenyi Biotec | 130-121-993 | DC |
| **DHR 123** | FITC | | Sigma-Aldrich | D1054 | GR |
| **HLA-DR** | VioGreen | REA805 | Miltenyi Biotec | 130-111-948 | DC |
| **Ki-67** | AlexaFluor700 | Ki-67 | BioLegend | 350530 | T1, T2 |
| **Viobility 405/452** | PB450/VioBlue | | Miltenyi Biotec | 130-110-205 | ID, T1,T2, GR, DC |

Abbreviations: ID = immune cell distribution panel, GR = granulocyte panel, DC = dendritic cells panel, T1 = T cell panel 1, T2 = T cell panel 2. All antibodies/dyes were used at 2% v/v, except for Ki-67 (5% v/v), DHR 123 (1% v/v), and Viobility 405/452 (1% v/v).
