## Supplementary figures and images for "COVID-19 patients share common, corticosteroid-independent features of impaired host immunity to pathogenic molds"

### Supplementary Figure 1

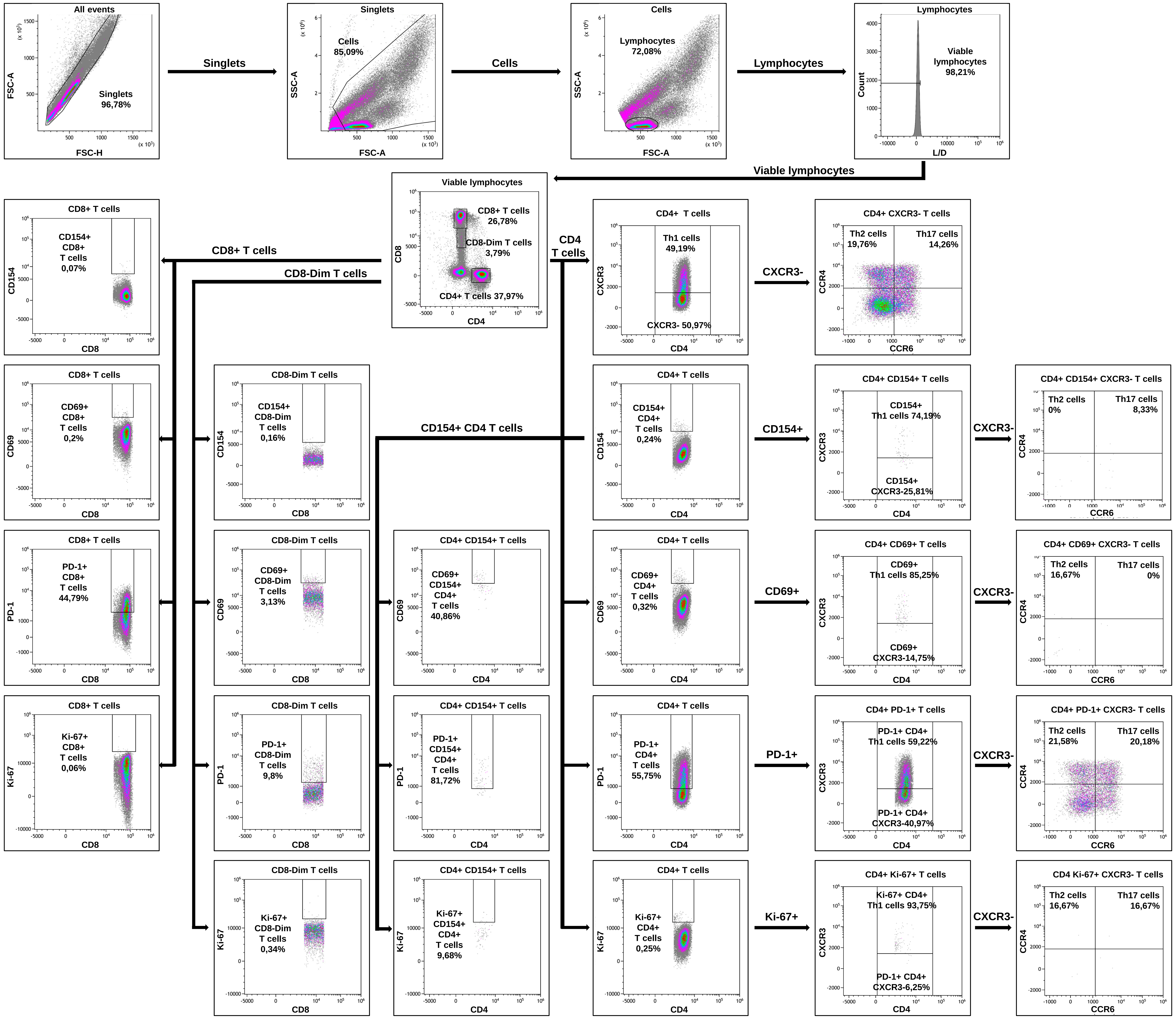

### Supplementary Figure 2

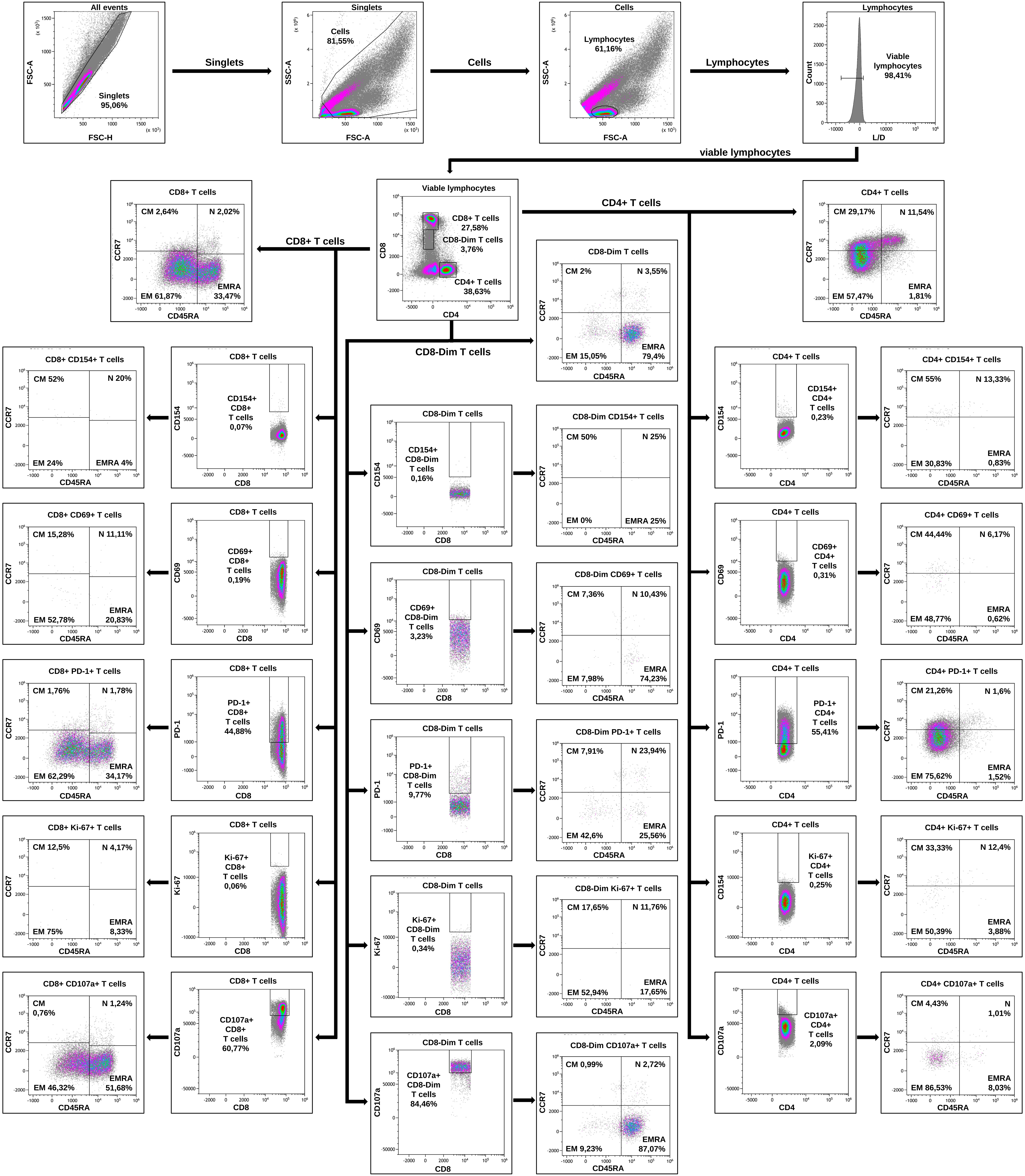

### Supplementary Figure 3

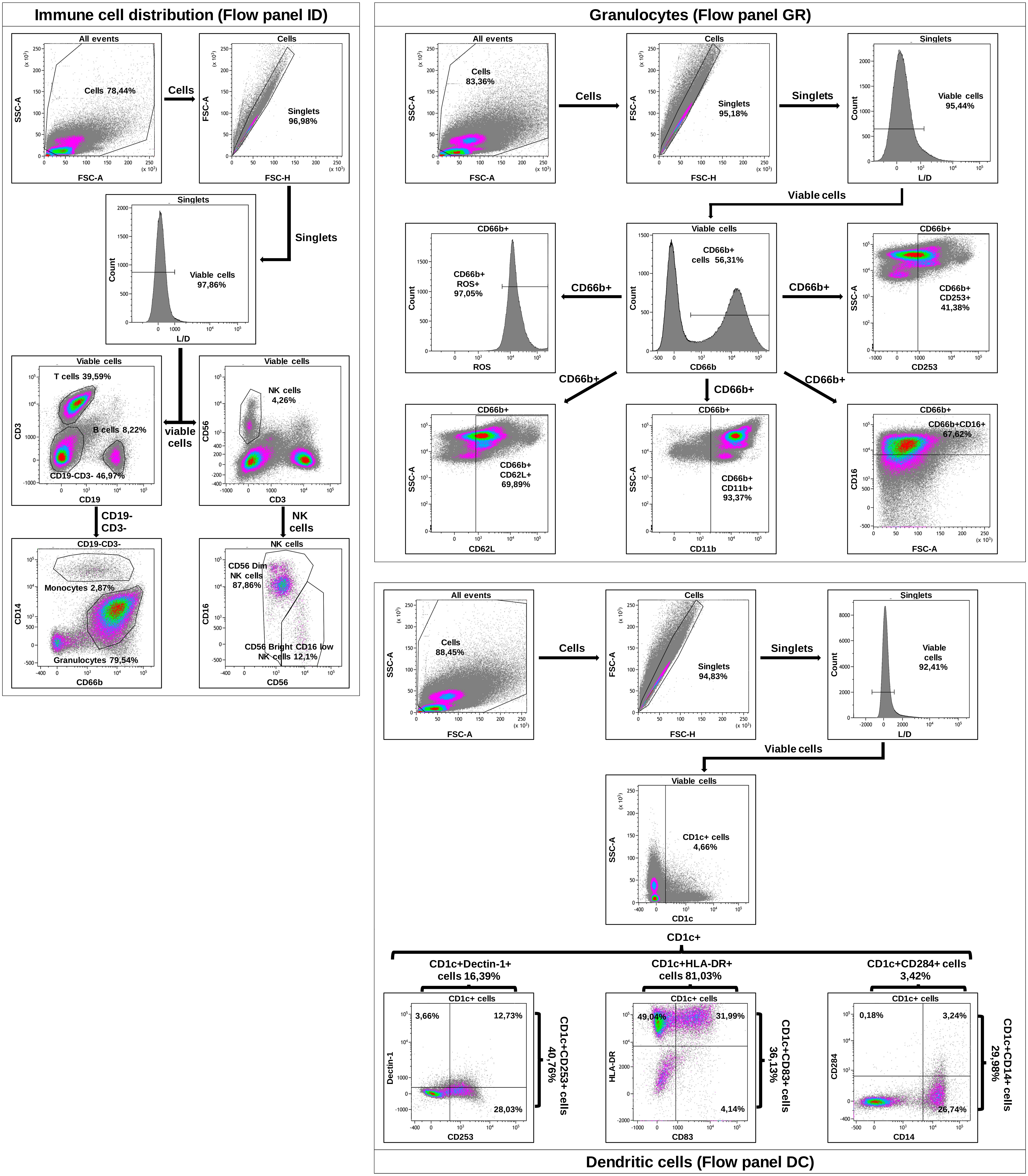

### Supplementary Figure 4

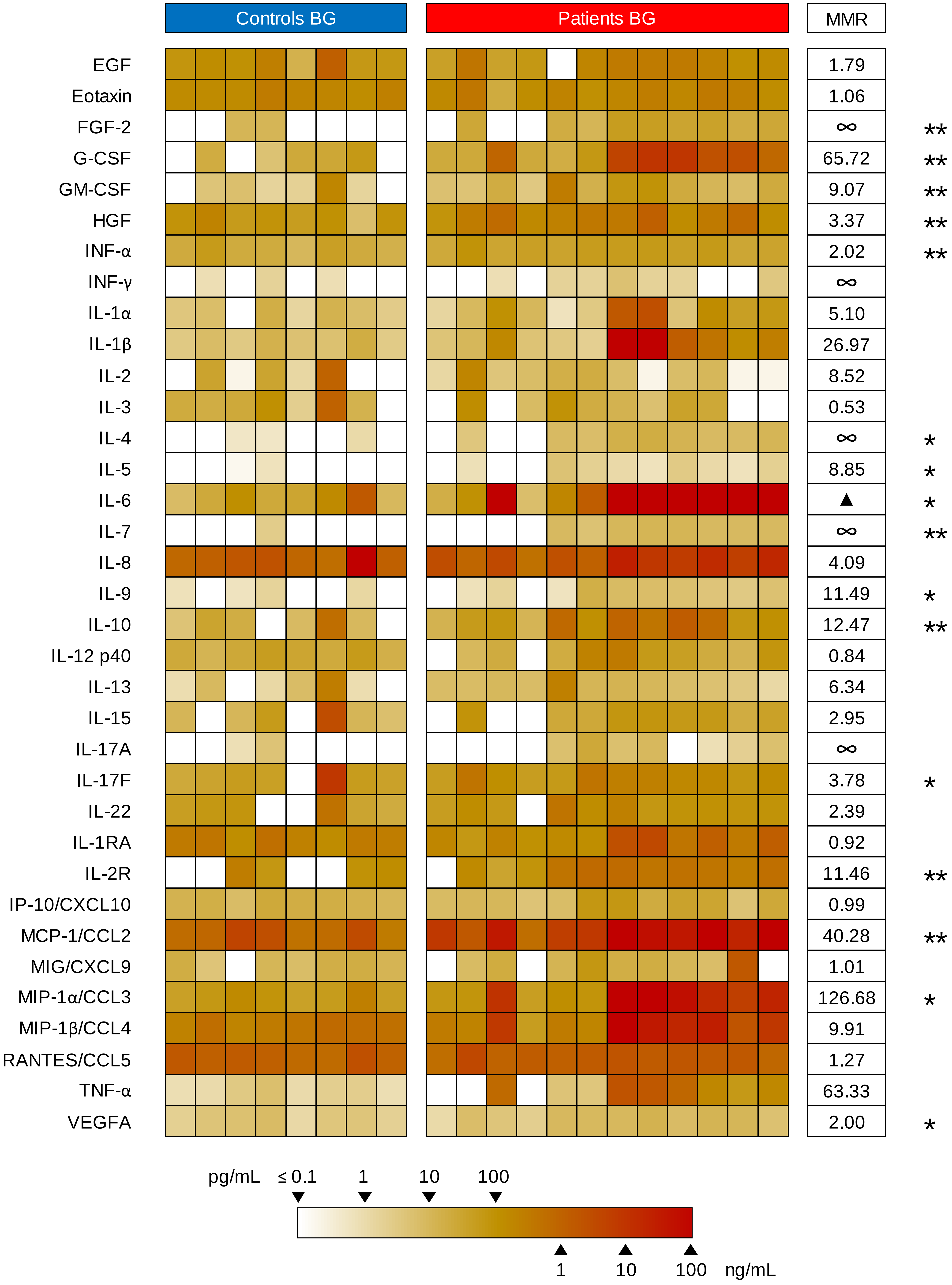

### Supplementary Figure 5

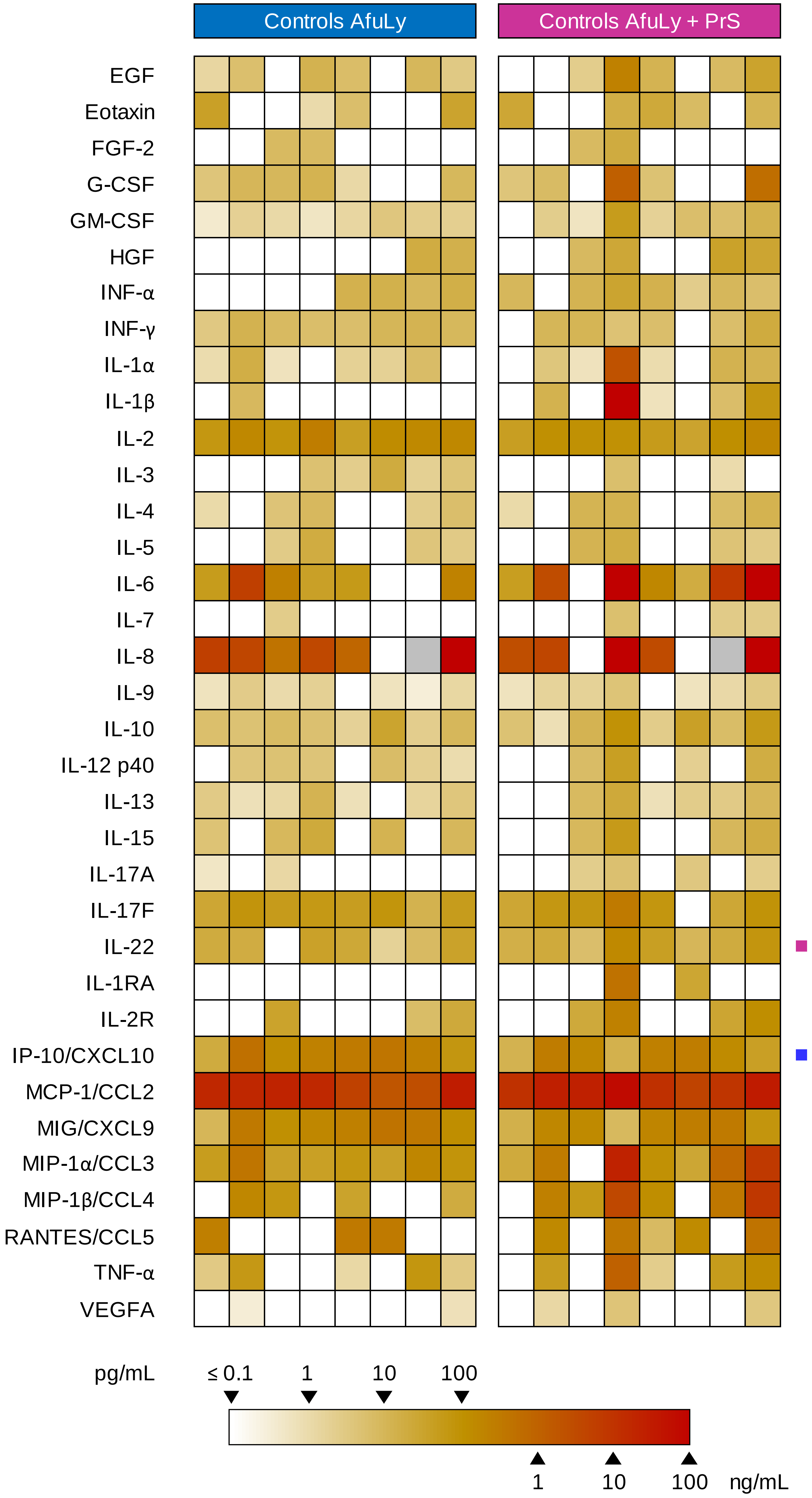

### Supplementary Figure 6

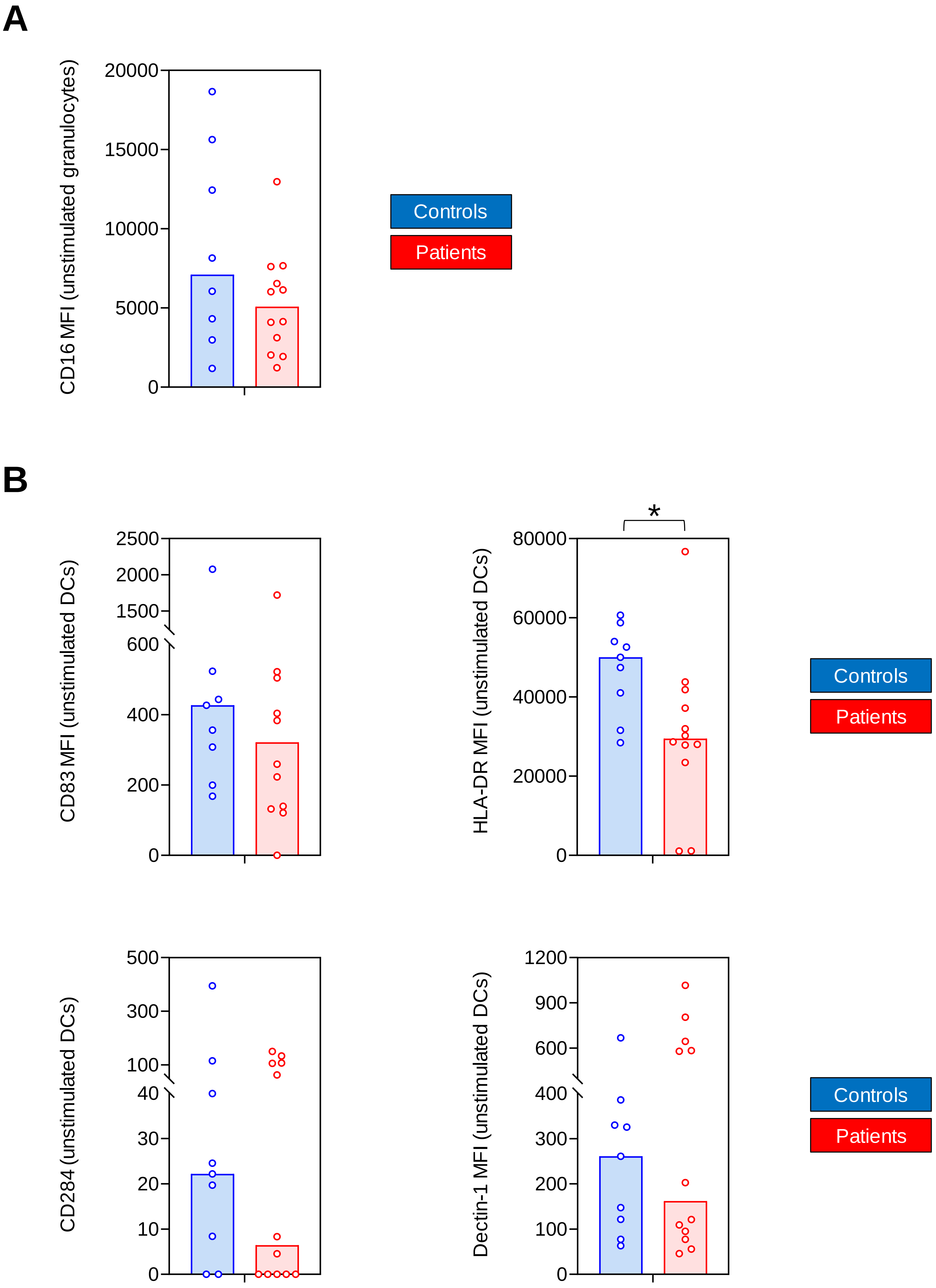

### Supplementary Figure 7

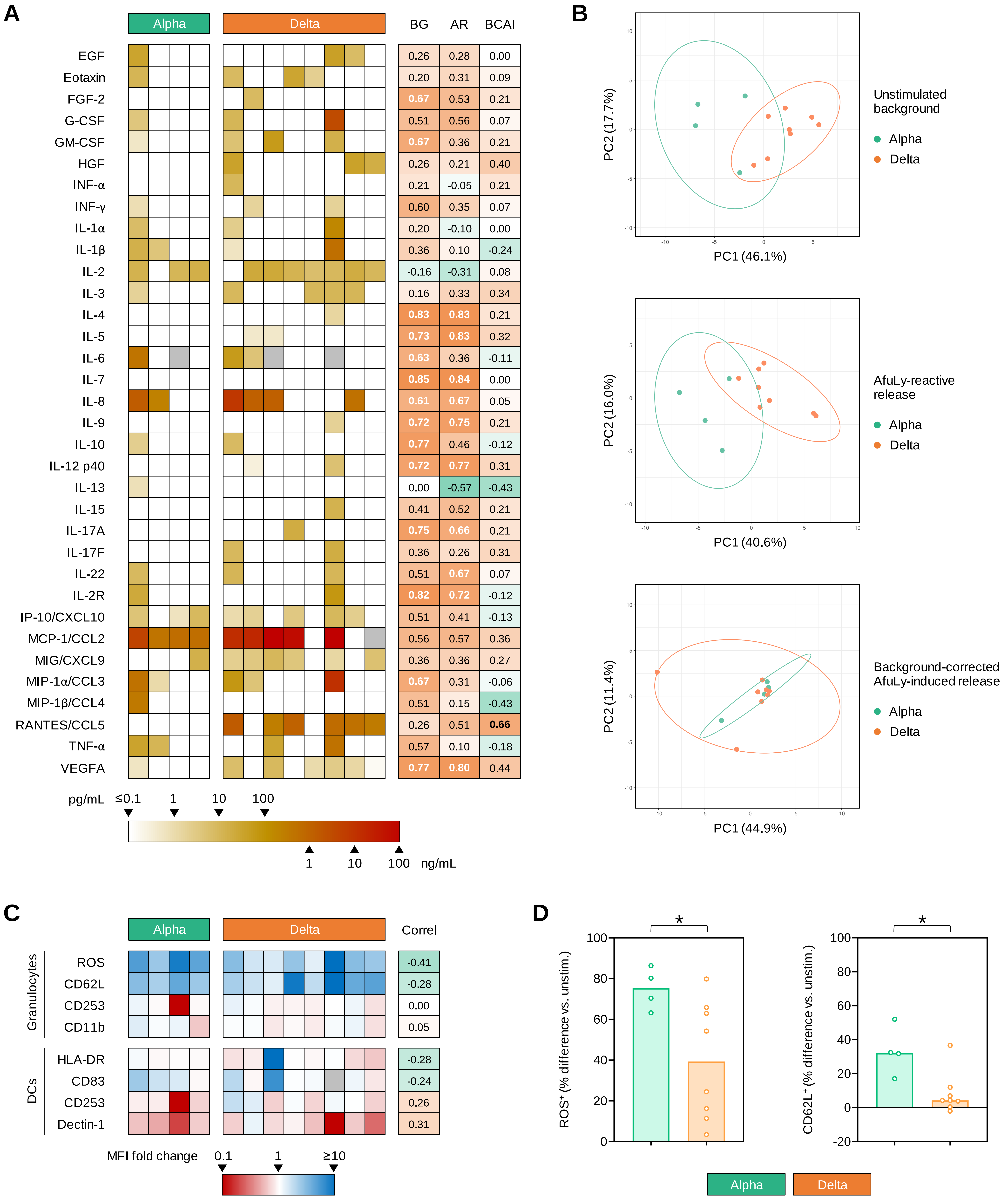

### Supplementary Figure 9

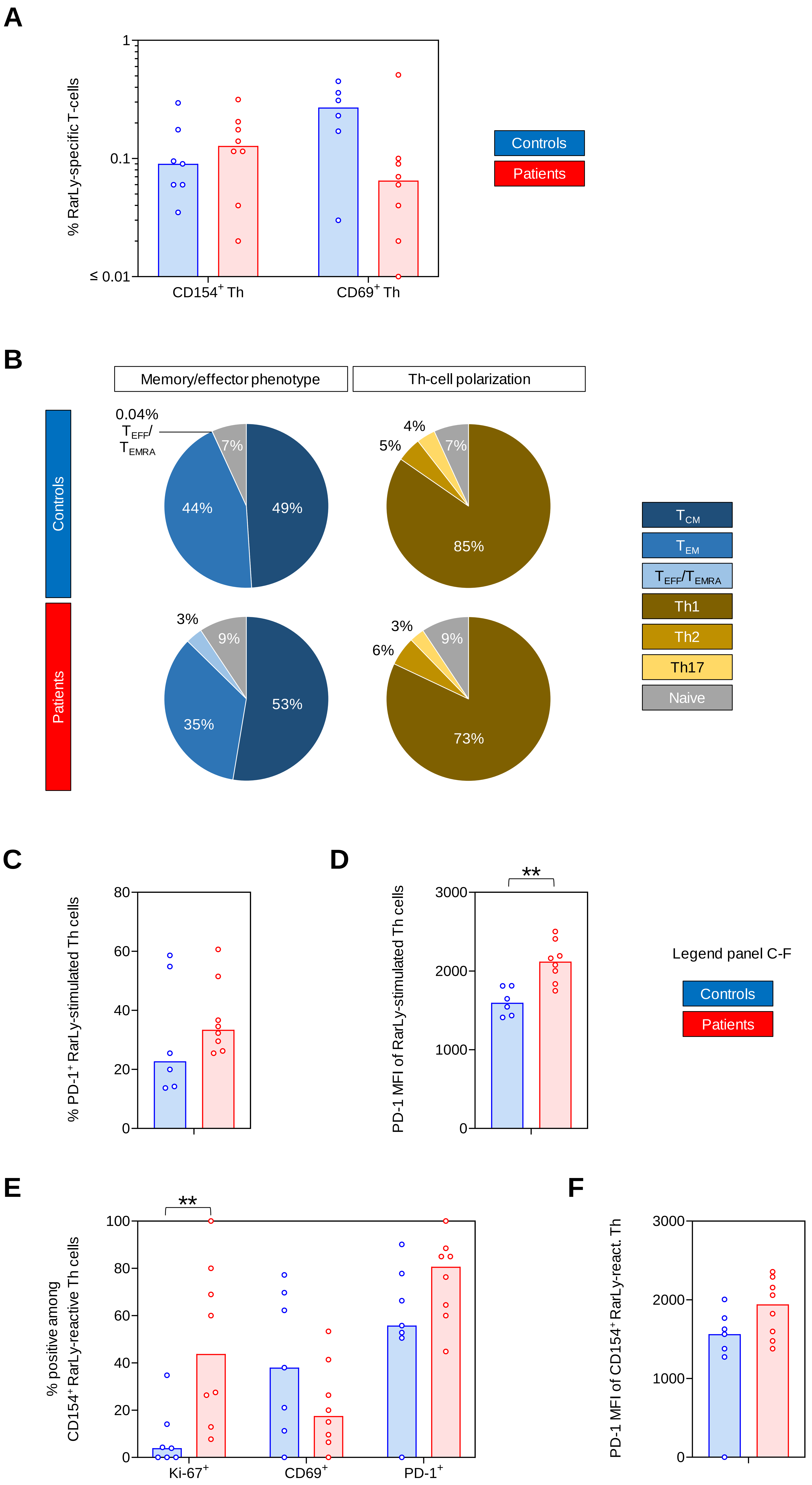
