## Supplementary Figure 8 for "COVID-19 patients share common, corticosteroid-independent features of impaired host immunity to pathogenic molds"

**A**

| % among AfuLy-stimulated Th | P vs. C Sign. | WHO | GCS | Time |
|---|---|---|---|---|
| CD154 | | 0.19 | -0.17 | 0.23 |
| CD69 | * | 0.31 | 0.18 | 0.23 |
| PD-1 | * | 0.09 | 0.37 | -0.01 |
| Ki-67 | | -0.37 | 0.00 | -0.02 |

Color scale for panels A, B, and D

ρ = -1 0 1

**B**

| | P vs. C Sign. | Max. WHO score BG | Max. WHO score AR | Max. WHO score BCAI | GCS dose BG | GCS dose AR | GCS dose BCAI | Time after pos. PCR BG | Time after pos. PCR AR | Time after pos. PCR BCAI |
|---|---|---|---|---|---|---|---|---|---|---|
| EGF | | -0.27 | -0.22 | -0.07 | -0.28 | -0.10 | 0.36 | 0.15 | 0.11 | -0.23 |
| Eotaxin | | **-0.63** | **-0.60** | -0.15 | 0.19 | 0.10 | -0.04 | -0.36 | -0.46 | -0.42 |
| FGF-2 | | -0.53 | -0.47 | 0.57 | -0.40 | -0.06 | -0.40 | -0.07 | 0.02 | 0.49 |
| G-CSF | | -0.49 | -0.30 | -0.12 | -0.47 | -0.20 | 0.46 | -0.10 | 0.06 | -0.48 |
| GM-CSF | | -0.39 | -0.10 | -0.25 | -0.40 | 0.13 | 0.34 | -0.19 | -0.12 | -0.18 |
| HGF | | -0.40 | -0.58 | -0.12 | -0.16 | 0.12 | -0.13 | 0.06 | -0.08 | -0.04 |
| INF-α | * | -0.22 | 0.12 | -0.21 | -0.21 | 0.12 | 0.18 | 0.06 | 0.14 | -0.40 |
| INF-γ | *** | -0.02 | 0.47 | 0.37 | **-0.60** | -0.17 | 0.09 | 0.27 | 0.18 | 0.08 |
| IL-1α | | -0.37 | -0.27 | -0.07 | -0.23 | 0.38 | 0.48 | 0.16 | 0.08 | -0.44 |
| IL-1β | | -0.56 | -0.51 | -0.15 | -0.28 | 0.32 | **0.66** | -0.15 | -0.05 | -0.39 |
| IL-2 | *** | 0.07 | 0.19 | 0.42 | 0.29 | 0.17 | -0.33 | -0.09 | 0.45 | **0.68** |
| IL-3 | | -0.27 | -0.40 | -0.37 | 0.10 | 0.09 | 0.20 | -0.34 | -0.45 | **-0.66** |
| IL-4 | ** | -0.43 | -0.36 | -0.21 | **-0.64** | -0.34 | 0.18 | -0.03 | 0.03 | -0.31 |
| IL-5 | * | -0.12 | 0.00 | 0.27 | -0.41 | -0.52 | -0.33 | -0.14 | 0.07 | 0.59 |
| IL-6 | | -0.53 | -0.27 | 0.29 | -0.37 | -0.01 | 0.36 | -0.15 | 0.11 | -0.05 |
| IL-7 | | -0.45 | -0.23 | 0.00 | **-0.62** | -0.42 | 0.00 | -0.20 | 0.00 | 0.00 |
| IL-8 | | -0.39 | -0.35 | 0.13 | -0.34 | -0.21 | 0.25 | -0.01 | -0.02 | 0.13 |
| IL-9 | *** | -0.07 | -0.27 | -0.21 | **-0.71** | -0.49 | 0.18 | 0.32 | 0.21 | -0.31 |
| IL-10 | *** | -0.49 | -0.34 | 0.04 | -0.35 | 0.18 | 0.42 | -0.35 | -0.33 | -0.32 |
| IL-12 p40 | * | -0.05 | 0.01 | 0.22 | **-0.66** | **-0.65** | -0.13 | 0.32 | 0.26 | 0.08 |
| IL-13 | *** | -0.11 | -0.24 | 0.31 | 0.09 | **0.78** | 0.40 | 0.04 | -0.47 | 0.00 |
| IL-15 | * | -0.48 | -0.37 | -0.21 | -0.25 | -0.19 | 0.18 | -0.03 | 0.08 | -0.31 |
| IL-17A | | 0.12 | 0.10 | -0.21 | **-0.68** | **-0.71** | -0.40 | 0.45 | 0.52 | 0.00 |
| IL-17F | *** | -0.16 | -0.27 | -0.31 | -0.35 | -0.05 | 0.26 | 0.27 | 0.21 | -0.52 |
| IL-22 | ** | -0.33 | -0.20 | -0.12 | -0.04 | -0.18 | 0.46 | -0.04 | 0.08 | -0.49 |
| IL-1RA | | -0.38 | -0.18 | 0.00 | -0.45 | -0.30 | 0.00 | -0.02 | 0.25 | 0.00 |
| IL-2R | | 0.07 | 0.08 | 0.04 | **-0.69** | -0.41 | 0.42 | 0.30 | 0.26 | -0.25 |
| IP-10/CXCL10 | *** | 0.00 | 0.32 | 0.26 | -0.44 | -0.55 | 0.11 | 0.22 | 0.40 | -0.07 |
| MCP-1/CCL2 | | -0.29 | -0.18 | 0.02 | -0.36 | -0.32 | -0.14 | 0.00 | 0.11 | 0.25 |
| MIG/CXCL9 | *** | -0.34 | -0.11 | 0.41 | -0.19 | -0.37 | -0.56 | -0.01 | 0.24 | 0.60 |
| MIP-1α/CCL3 | * | -0.54 | -0.43 | 0.12 | -0.48 | 0.14 | 0.44 | -0.14 | -0.16 | -0.22 |
| MIP-1β/CCL4 | | -0.58 | -0.43 | 0.31 | -0.22 | 0.37 | 0.40 | -0.20 | -0.08 | 0.00 |
| RANTES/CCL5 | | -0.47 | **-0.60** | -0.44 | -0.06 | -0.14 | -0.22 | -0.07 | -0.22 | -0.22 |
| TNF-α | | -0.57 | -0.47 | -0.15 | -0.36 | 0.29 | 0.53 | -0.20 | -0.05 | -0.06 |
| VEGFA | | -0.55 | **-0.59** | -0.33 | -0.36 | -0.23 | 0.11 | -0.16 | -0.22 | -0.29 |

**C**

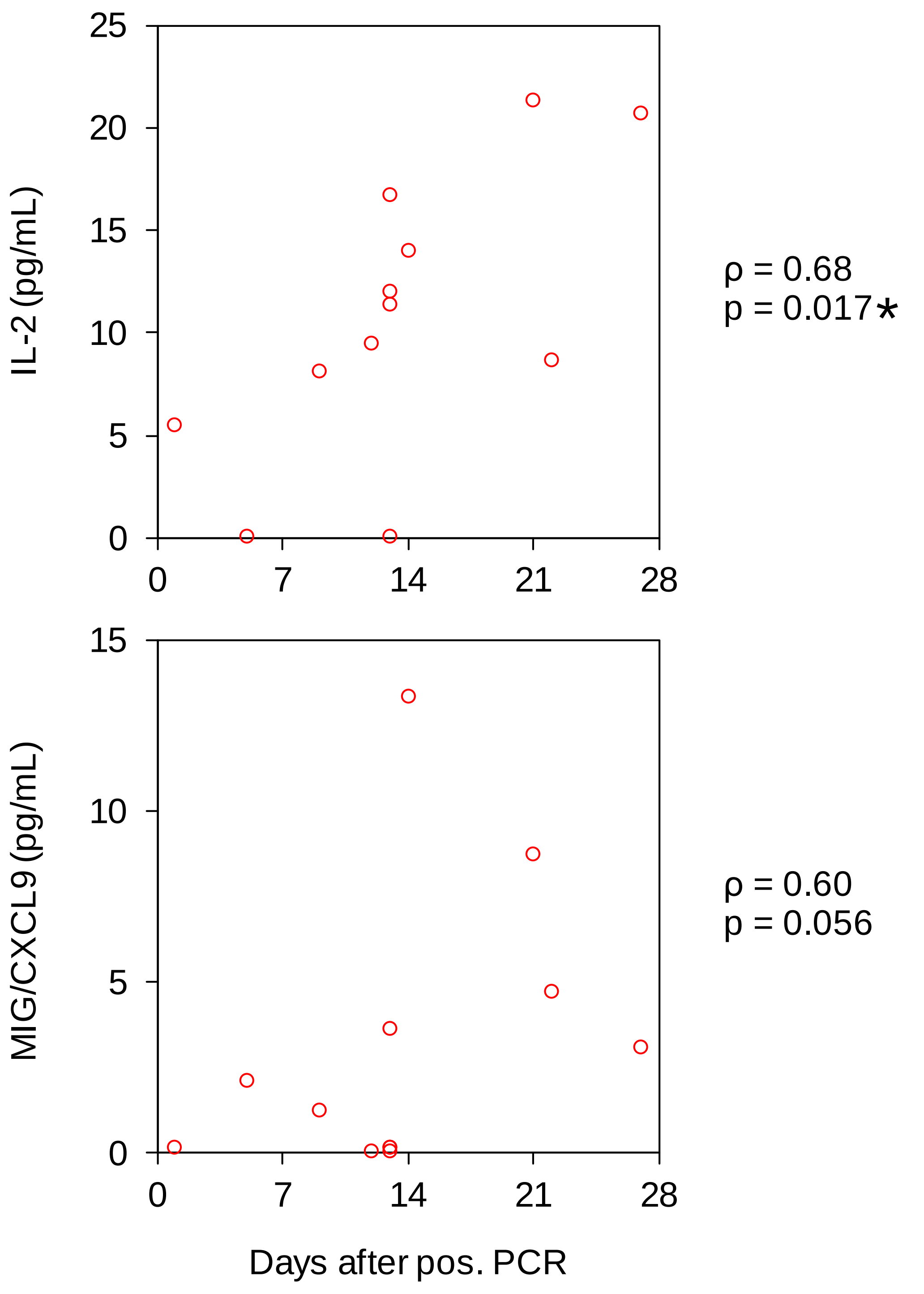


**D**

| | | | P vs. C Sign. | WHO | GCS | Time |
|---|---|---|---|---|---|---|
| Granulocytes | % pos. | ROS | *** | -0.17 | 0.72 | -0.26 |
| | | CD62L | *** | 0.30 | 0.47 | 0.01 |
| | | CD253 | * | -0.16 | 0.57 | -0.48 |
| | | CD11b | | -0.12 | 0.17 | -0.03 |
| | MFI | ROS | *** | 0.00 | 0.56 | -0.30 |
| | | CD62L | ** | -0.14 | 0.04 | -0.21 |
| | | CD253 | ** | 0.13 | 0.25 | -0.13 |
| | | CD11b | | -0.23 | 0.62 | -0.33 |
| DCs | MFI | HLA-DR | | -0.17 | 0.35 | -0.09 |
| | | CD83 | | -0.53 | 0.75 | -0.40 |
| | | CD253 | | -0.14 | 0.09 | -0.35 |
| | | Dectin-1 | | 0.26 | -0.49 | 0.41 |

**E**

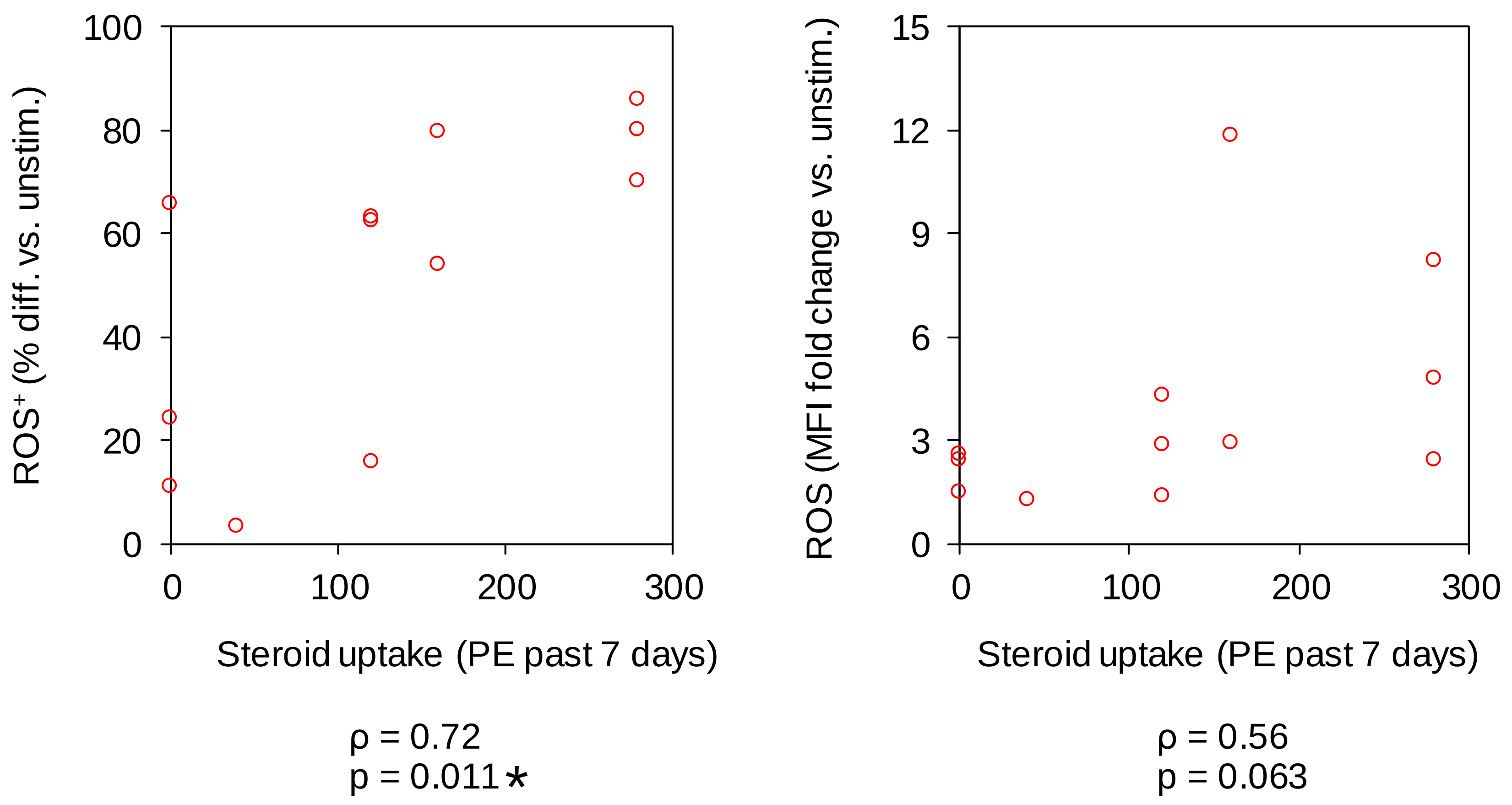
